# The Genetic Architecture of Cervical Length is Shared with Spontaneous Preterm Birth Risk

**DOI:** 10.1101/2024.05.23.24307808

**Authors:** Hope M. Wolf, Bradley T. Webb, Jerome F. Strauss, Adi L. Tarca, Roberto Romero, Sonia S. Hassan, Shawn J. Latendresse, Tinnakorn Chaiworapongsa, Stanley Berry, Nardhy Gomez-Lopez, Piya Chaemsaithong, Timothy P. York

## Abstract

**Study Question:** Is there a genetic contribution to the change in cervical length during pregnancy and, if so, how is this related to the known genetic contribution to pregnancy duration?

**Summary Answer:** Genomic analyses suggest that cervical length change across pregnancy is a polygenic trait with appreciable heritability. Bivariate genetic correlations estimated using genome-wide complex trait analysis (GCTA) indicated that a large proportion of the genes influencing cervical change across pregnancy also influenced gestational duration.

**What is Known Already:** Sonographic cervical length is a powerful predictor of maternal risk for spontaneous preterm birth (sPTB). Twin and family studies have established a maternal genetic heritability for sPTB ranging from 13 - 20%. However, there is no corresponding estimate for the heritability of mid-trimester cervical length, or an understanding of how genetic factors contribute to cervical changes across pregnancy.

**Study Design, Size, Duration:** This study was based on a prospective longitudinal cohort of (N = 5,160) Black/African American women who underwent serial sonographic examination of the uterine cervix during pregnancy and were followed until delivery. Maternal DNA extracted from whole-blood samples was genotyped via next-generation low-pass whole genome sequencing.

**Participants/Materials, Settings, Methods:** Repeated measurements of sonographic cervical length were collected during singleton pregnancies in 5,160 unique women and longitudinal changes in cervical length were described using latent growth models. Individual-level low-pass whole genome sequencing data from a subset of 4,423 women were used to calculate the heritability of cervical length change during pregnancy and estimate its genetic correlation with gestational duration.

**Main Results and the Role of Chance:** Changes in cervical length across pregnancy were found to be substantially influenced by genetic factors (*h^2^* = 51%). Furthermore, the genetic factors influencing cervical change were significantly correlated with genetic influences on pregnancy duration, implying a shared genetic architecture. The strongest genetic correlation was observed for the overall linear change in cervical length (*r_g_* = 0.982 [95% CI 0.951, 0.993]), and less so for the genetic correlation with mid-trimester values (*r_g_* = 0.714 [95% CI 0.419, 0.873]) and aspects of non-linear change that are most pronounced at the end of pregnancy (*r_g_*= -0.490 [95% CI -0.766, -0.062]). SNP-level associations were observed near genes involved in the progesterone, estrogen, and insulin signaling pathways.

**Limitations, Reasons for Caution:** While the study cohort was not designed to identify individual genetic variants associated with cervical length change or gestational duration, it was well powered to assess aggregate genome-wide summary statistics to estimate trait heritability, bivariate genetic correlations, and genetic enrichment of suggestive associations. To date, this study is the largest genome-wide study of preterm birth in a cohort of Black/African American women, a population that is disproportionately affected by health disparities in preterm birth and perinatal outcomes.

**Wider Implications of the Findings:** The significant genetic correlation between cervical length and gestational duration suggests shared causal loci between these two traits. These results raise the question of whether a large proportion of genetic loci for gestational duration exert their influence through the process of cervical remodeling. Polygenic profiling of maternal genetic liability to cervical shortening could aid in the development of clinical risk assessment tools to identify high-risk women who may benefit from more frequent cervical length screening and earlier interventions to prevent preterm delivery.

**Study Funding/Completing Interest(s):** This research was supported, in part, by the Perinatology Research Branch, Division of Obstetrics and Maternal-Fetal Medicine, Division of Intramural Research, *Eunice Kennedy Shriver* National Institute of Child Health and Human Development, National Institutes of Health, United States Department of Health and Human Services (NICHD/NIH/DHHS); and, in part, by federal funds from NICHD/NIH/DHHS (Contract No. HHSN275201300006C). RR has contributed to this work as part of his official duties as an employee of the United States Federal Government. ALT, NGL, and SSH were also supported by the Wayne State University Perinatal Initiative in Maternal, Perinatal and Child Health. Additional support was provided from the March of Dimes Prematurity Research Center at the University of Pennsylvania (22-FY18-812).

**Trial Registration Number:** N/A.

## INTRODUCTION

The cervix undergoes carefully orchestrated compositional changes throughout pregnancy, transitioning from a firm structure that protects the *in utero* environment during pregnancy to a soft and pliable tissue allowing for parturition. In most pregnancies delivered at term, a gradual decrease in cervical length (CL) is observed around 30 weeks of gestation, shortening from approximately 35-45 mm to complete effacement by the first stage of labor. However, this process of cervical shortening begins earlier or happens more rapidly in some pregnancies. A short cervix is defined as a sonographic cervical length of less than 25 mm before 24 weeks of gestation, and is associated with a 6-fold increase in the risk for preterm birth (i.e., birth before 37 completed weeks)(Iams *et al*., 1996). The faster cervix shortens, and the earlier in pregnancy that shortening begins, the higher the risk for spontaneous preterm delivery.

There is a clear pathophysiological connection between cervical dysfunction and spontaneous preterm birth, and several maternal risk factors are associated with both conditions, including maternal age, parity, pre-pregnancy BMI, race, and previous abortion (Goldenberg *et al*., 2008; Anum *et al*., 2010; Buck *et al*., 2017; Cho *et al*., 2017; Kandil *et al*., 2017; Gudicha *et al*., 2021; Brittain *et al*., 2023). A recent study showed that many maternal risk factors for preterm birth are mediated through their effects on cervical change during pregnancy (Wolf *et al*., 2023). Yet, whether the previously estimated genetic influences on spontaneous preterm birth (Lunde *et al*., 2007; Svensson *et al*., 2009; York *et al*., 2010, 2013, 2014) are related to or operate through mechanisms that affect cervical length remains unclear. While genome-wide association studies (GWAS) have identified a handful of loci associated with gestational duration (Zhang *et al*., 2017; Solé-Navais *et al*., 2023), there have been no reported heritability estimates of cervical length and only limited efforts to identify individual loci associated with cervical dysfunction (Volozonoka *et al*., 2020).

Meta-analyses of twin and family studies have established that nearly all human complex traits are influenced by genetic factors (Claussnitzer *et al*., 2020). An average heritability of 45.1% was observed across 64 traits in heritability studies specifically related to female reproduction, indicating that nearly half of interindividual variation in these traits could be attributed to genetic factors (Polderman *et al*., 2015; Claussnitzer *et al*., 2020). Estimates for the heritability of spontaneous preterm birth range from 14 – 40% based on twin and family studies (Wadon *et al*., 2020) and 13 – 21% based on single nucleotide polymorphism (SNP) genotype data (York *et al*., 2014; Zhang *et al*., 2017). Heritability estimates are often correlated within functional domains, suggesting that traits related to a particular biological function share similar genetic influences. The extent to which changes in cervical length and gestational duration share a genetic architecture (i.e.,the number, location, frequency, interactions, and effect sizes of genetic variants influencing each trait) can be quantified by calculating their bivariate genetic correlation. A better understanding of the genetic and mechanistic relationships between cervical changes during pregnancy and the risk of gestational duration is needed to make progress in predicting, preventing, and treating adverse maternal and neonatal outcomes associated with premature delivery.

Since there is currently no characterization of the genetic architecture of cervical length during pregnancy or how these genetic factors relate to gestational duration, this study focused on the following fundamental questions: What aspects of cervical length change across pregnancy are heritable (i.e., mid-pregnancy length, linear, and nonlinear change components)? To what extent do genetic influences on cervical change reflect a polygenic pattern of inheritance? And how much of this genetic contribution, if any, is shared with genes affecting gestational duration?

Heritability estimates and the genetic correlations among traits were estimated by conducting a GWAS alongside Genome-Wide Complex Trait Analysis (GCTA) of low-pass whole-genome sequencing data in a cohort of pregnant Black/African American women with serial cervical length measurements obtained across pregnancy. Additional insight into the biological mechanisms underlying cervical change during pregnancy was gained by integrating GWAS results with functional and pathway annotations, including tissue-specific gene expression data, epigenetic marks, and chromatin accessibility.

## MATERIALS AND METHODS

### Cohort Description and Phenotyping

The genotyped cohort was composed of self-identified Black/African-American women from the Detroit, Michigan area enrolled in a prospective study of pregnant women. Serial sonographic CL measurements between 8 - 40 weeks of gestation were available for 5,160 women carrying singleton pregnancies. In this study, gestational duration was evaluated as both gestational age at delivery (GAD) and spontaneous preterm birth (sPTB). GAD was measured in weeks from the first day of a woman’s last menstrual period (confirmed by ultrasound) to the day of delivery. sPTB was defined as an uninduced delivery before 37 completed weeks of gestation, including births following both spontaneous preterm labor and preterm premature rupture of membranes (PPROM). Data from patients treated with progesterone or who had cerclage (n=260) were excluded from the analysis as these interventions may alter the natural progression of cervical shortening during pregnancy. Cervical change across gestation was quantified for each pregnancy in a previous study using Latent Growth Curve Analysis (LGCA) (Wolf *et al*., 2022, 2023), a cross-disciplinary analytic approach that leverages repeated measurements to estimate how a trait changes within and between individuals over time. Interindividual differences in cervical change during pregnancy were described by coefficients of the growth curve corresponding to the model intercept, linear change (*i.e.*, slope), and nonlinear (*i.e.*, quadratic) change (**Figure 1**). The model intercept was centered at 17-20 weeks so that the estimated value of this coefficient would correspond to the midtrimester cervical length (MTCL). The cervical change coefficients and gestational duration measures (*i.e.*, GAD, sPTB) were used as traits in the GCTA and GWAS analyses.

**Figure 1.**
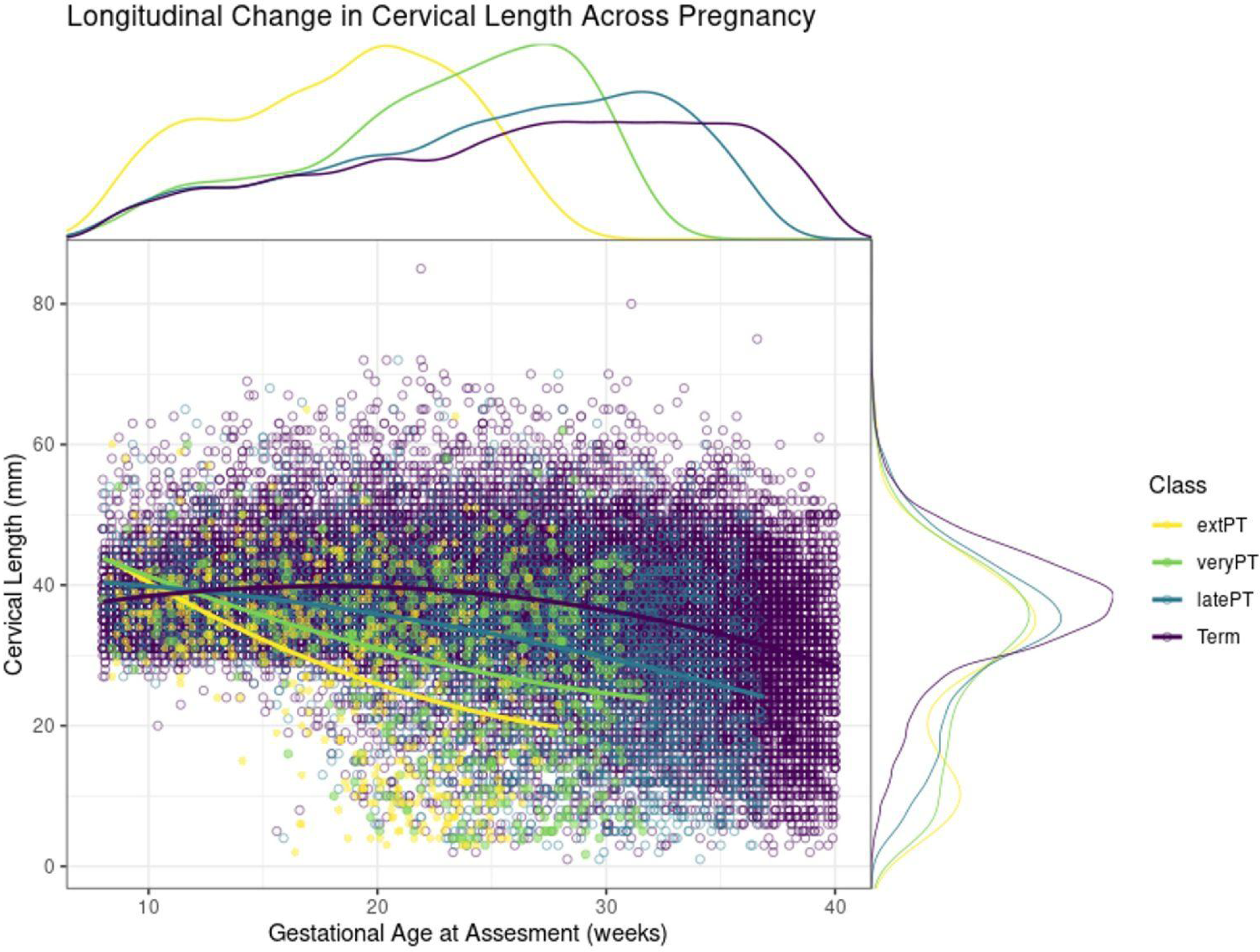
**A.** Serial measurements of CL within individuals across pregnancy. Marginal plots show distribution of CL data across gestational ages and birth outcome classes. Average trajectories of CL change during pregnancy estimated by LGCA are shown for four classes of clinical birth outcomes (extremely preterm births delivered before 28 weeks gestation, very preterm births delivered before 32 weeks gestation, late preterm births delivered before 37 weeks gestation, and full term births delivered after 37 completed weeks of gestation) are shown in color. The trajectory and rate of cervical change during pregnancy varies by gestational age at delivery.

### Ethical Approval

Participants were enrolled under the protocols for Biological Markers of Disease in the Prediction of Preterm Delivery and for Preeclampsia and Intra-Uterine Growth Restriction: A Longitudinal Study (WSU IRB#110605MP2F and NICHD/NIH# OH97-CH-N067) between 2005 – 2017 at the Center for Advanced Obstetrical Care and Research (CAOCR) at Hutzel Women’s Hospital. The Institutional Review Boards of Wayne State University and the *Eunice Kennedy Shriver* National Institute of Child Health and Human Development (NICHD)/National Institutes of Health (NIH)/U.S. Department of Health and Human Services (DHHS) (Detroit, MI, USA) approved the study. All participants provided written informed consent for the collection of cervical length data and blood samples for future genetic research studies.

### Whole-Genome Sequencing and Quality Control

Purified genomic DNA (gDNA) was extracted from buffy coat samples and genotyped by BGI Americas Corporation using paired-end low-pass, whole genome sequencing. Gencove, Inc. performed the sequence alignment using the GRCh37-hg19 human reference genome assembly, variant calling, phasing, and imputation based on the loimpute v0.18 algorithm for low-pass sequencing data (Li *et al*., 2021). A recombination map derived from the HapMap II project was used to interpolate recombination rates across all sites in the imputation reference panel. After imputation, genetic variant information was available for 65 million SNPs.

The mean effective sequencing depth coverage of the sample was 1.45x. Variants with a genotyping rate of <5% (geno < 0.05); a MAF of <0.5% (MAF < 0.005); or that were found to be in extreme deviation from Hardy-Weinberg equilibrium (HWE < 0.000001) were excluded. Approximately 19.8 million single-nucleotide polymorphisms (SNPs) remained after these quality control filters. A principal component analysis (PCA) of genotypes was conducted in Plink (Purcell *et al*., 2007; Chang *et al*., 2015) based on an external reference panel of known ancestry reference (1000 Genomes Project, Phase 3) (1000 Genomes Project Consortium *et al*., 2015). Samples were excluded due to poor quality (n=2); missing data (n=15); duplicates (n=80); and population stratification concerns (n=243). In total, 4423 samples passed all quality control measures for genetic analyses (**Figure 2**).

**Figure 2.**
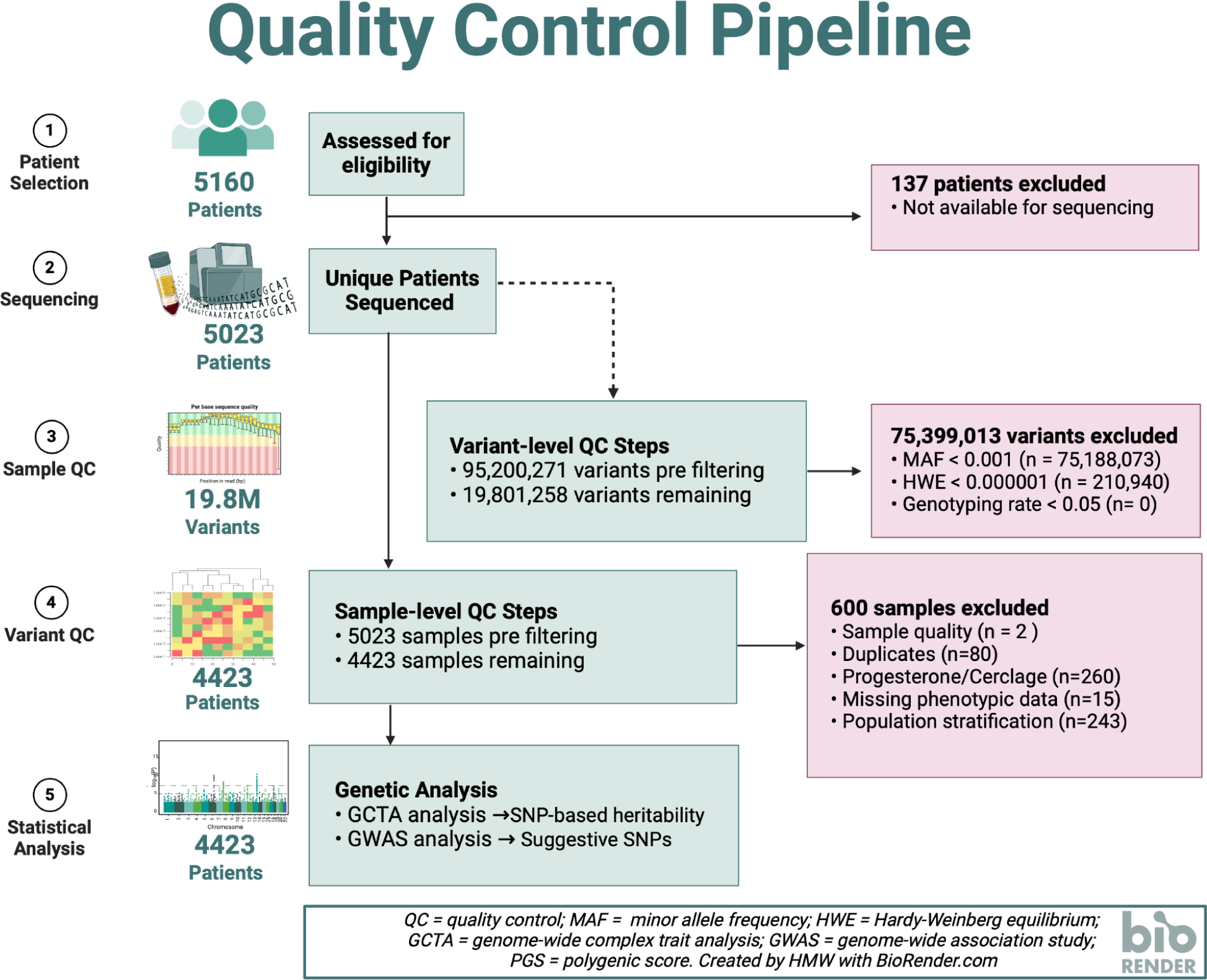
Quality control pipeline for genomic analyses. Purified genomic DNA from 5023 patients with available samples were sent for low-pass whole-genome sequencing. Approximately 9.8 million single-nucleotide polymorphisms (SNPs) remained after variant level filtering for genotyping rate (geno < 0.05), minor allele frequency (MAF < 0.005), and deviation from Hardy-Weinberg equilibrium (HWE < 0.000001). Samples from 4423 women remained after excluding samples for poor nucleic acid quality (n=2), missing data (n=15), duplicates (n=80), and population stratification (n=243).

### Trait Heritability and Bivariate Genetic Correlation Analysis

The SNP-based heritability of cervical change coefficients was estimated using restricted maximum likelihood (REML) as implemented in GCTA version 1.92.3 (Yang *et al*., 2011). Covariates in the heritability analysis included sequencing depth, batch number, maternal height, pre-pregnancy BMI, parity, self-reported race/ethnicity, and the first 10 SNP-based ancestry principal components (PCs). Both self-reported race/ethnicity and ancestry principal components were included as covariates as they capture different aspects of environmental and genetic factors that may account for trait variance. SNP-based heritability was estimated for maternal height, pre-pregnancy BMI, GAD, and sPTB to compare the within-cohort estimates to SNP and twin-based heritability published in the literature. Bivariate GCTA was used to decompose observed correlations between traits into covariance components explained by genetic and environmental factors. Bivariate GCTA methods were used to estimate the genetic correlation between the cervical change coefficients and gestational duration measures.

### Genome-Wide Associations

Although identification of an appreciable set of individual SNP associations was not deemed practical given the available sample size (Visscher *et al*., 2017; Wu *et al*., 2022), an exploratory analysis of genome-wide associations was conducted to gain insight into potential genetic mechanisms and biological pathways associated with cervical change and pregnancy duration. The whole genome association analysis toolset Plink (PLINK v2.00a3.3LM 64-bit Intel)(Purcell *et al*., 2007; Chang *et al*., 2015) was used to test for genetic associations with cervical change coefficients and gestational duration. PLINK dosages for the X chromosome in the all-female samples were set to levels similar to regular diploid chromosomes. Generalized linear models were used to test associations for the quantitative traits (cervical length coefficients, and GAD), while a logistic regression model was used for sPTB. Sequencing depth, batch number, self-reported race/ethnicity, height, pre-pregnancy BMI, parity, and the first 10 PCs from the PCA were variance-standardized and included as covariates. Due to the shorter extent of linkage disequilibrium (LD) in populations with African ancestry, individual SNP associations in this cohort were evaluated using the more conservative threshold for genome-wide significance (p < 1.6×10^−8^) based on a Bonferroni correction for an estimated 3.0 million independent LD blocks.

### Annotation of GWAS Findings

The integrative web-based platform FUMA (Watanabe *et al*., 2017) was used to annotate variant-level summary statistics from suggestive loci (p < 1.0×10^−5^) associated with cervical change coefficients and gestational duration outcomes. Functional annotation and the mapping of SNPs to nearby genes was performed using the SNP2GENE function, while GENE2FUNC was used to obtain insights into putative biological mechanisms of mapped genes. Tissue specific expression patterns for the genes mapped by SNP2GENE were assessed in 53 tissue types using RNA-seq data from the Adult Genotype Tissue Expression (GTEx) v8 (GTEx Consortium, 2020). Gene-wise associations (de Leeuw *et al*., 2015) and Multi-marker Analysis of GenoMic Annotation (MAGMA) of the combined set of loci associated with the cervical change coefficients was used for exploratory gene-set enrichment analyses (de Leeuw *et al*., 2015) for a comprehensive test of genetic effects on cervical length change.

### Pre-registration

Analyses were pre-registered with the Center for Open Science using the AsPredicted format (https://osf.io/km2cg).

## RESULTS

### SNP-Based Heritability Estimates for Cervical Length Change

Cervical change during pregnancy was found to be highly heritable in the filtered sample of 4,423 women retained for genetic analyses (**Table 1**). Genetic sources explained at least 50% of inter-individual trait differences in cervical change during pregnancy (*h*^2^ = 0.509 [95% CI 0.327, 0.691]; *h*^2^ = 0.509 [95% CI 0.323, 0.695]; *h*^2^ = 0.507 [95% CI 0.325, 0.689]) (**Figure 3, Table S1**). The estimated heritability of GAD (*h*^2^ = 0.158 [95% CI -0.022, 0.338) and spontaneous preterm birth (*h*^2^ = 0.154 [95% CI -0.024, 0.332) were consistent with previously reported values obtained by twin and family methods (York *et al*., 2014; Zhang *et al*., 2017). For instance, the SNP-based heritability estimated for GAD in this cohort was remarkably similar to the only twin and family study (York *et al*., 2010) that has estimated maternal heritability for GAD in African American women (i.e., 15.8% vs. 13.8%). Heritability estimates for height (*h*^2^ = 0.675 [95% CI 0.506, 0.844]) and BMI (*h*^2^ = 0.576 [95% CI 0.400, 0.752]) were also consistent with published estimates for these traits in other cohorts (Guo *et al*., 2021; Yengo *et al*., 2022). These results show that cervical changes during pregnancy are highly heritable, and that heritability estimates from this cohort align with those reported in other studies.

**Figure 3.**
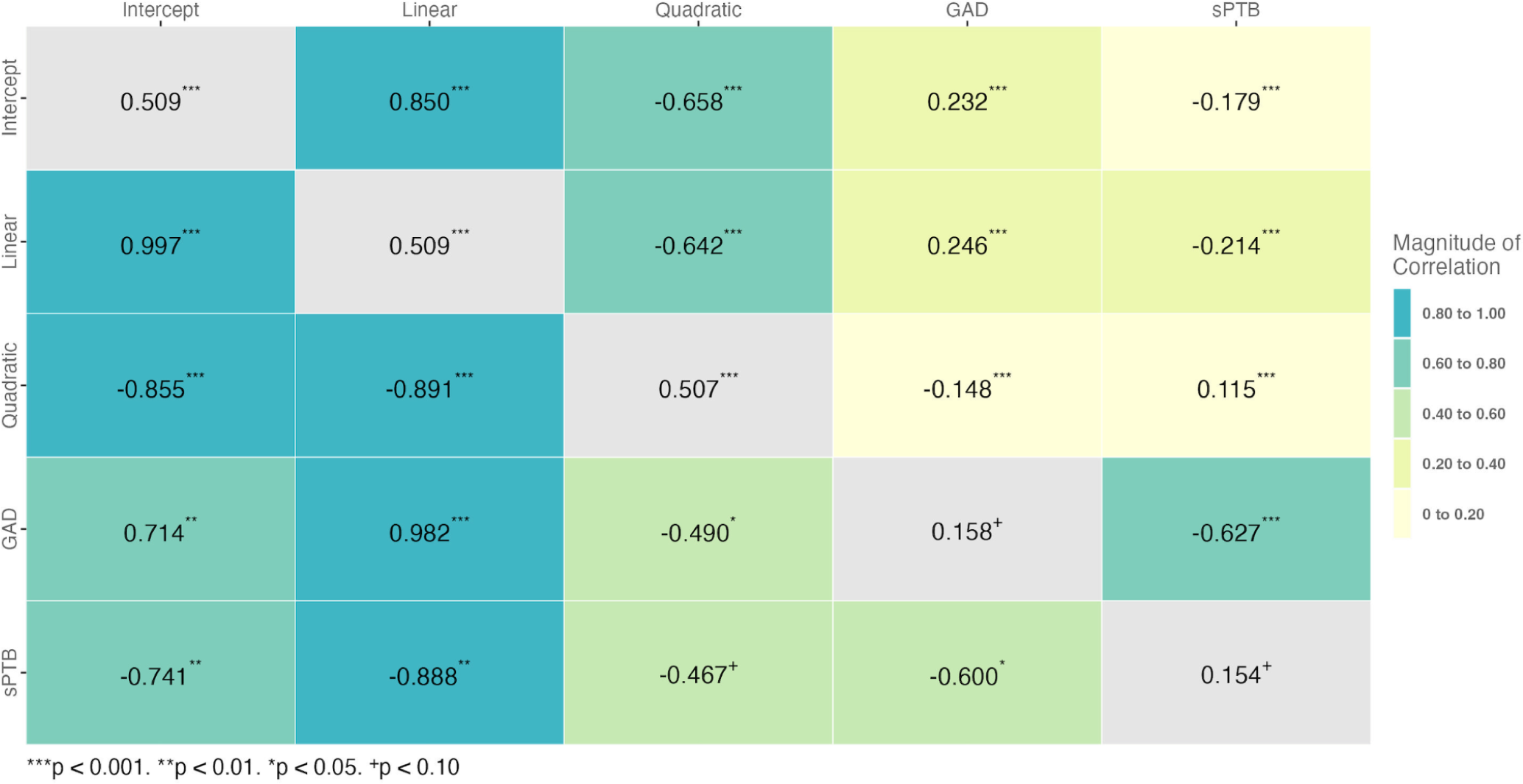
Matrix of heritability estimates and genetic and trait correlations estimated between trait pairs in the cohort. Values in the upper triangle represent observed phenotypic correlations between traits, while values in the lower triangle represent genetic correlations between traits. Values along the diagonal represent a trait’s SNP-based heritability. Note that negative genetic correlations occur when an increase in trait 1 corresponds with a decrease in trait 2.

**Table 1.** Cohort Demographic Table for the n = 4423 women with low-pass whole-genome sequencing data who were included in the GCTA and GWAS genetic analyses. MTCL = observed midtrimester cervical length measurement, sCL = short cervix (defined as CL < 25 mm before 24 weeks gestation).

| Cohort Demographic Table |  |  |  |  |
| --- | --- | --- | --- | --- |
| Variable | Overall, N = 4,422 | Preterm, N = 563 | Term, N = 3,859 | p-value<sup>1</sup> |
| **Age, Median (IQR)** | 23.0 (20.0 – 27.0) | 23.0 (20.0 – 28.0) | 23.0 (20.0 – 27.0) | 0.036 |
| **Self Reported Race/Ethnicity, n (%)** |  |  |  | 0.31 |
| Black | 4,127 (93) | 531 (94) | 3,596 (93) |  |
| Other | 295 (6.7) | 32 (5.7) | 263 (6.8) |  |
| **Maternal Height (in), Median (IQR)** | 64.00 (62.00 – 66.00) | 64.00 (62.00 – 66.00) | 64.00 (62.00 – 66.00) | 0.21 |
| **Pre-Pregnancy BMI, Median (IQR)** | 27 (23 – 33) | 27 (22 – 33) | 27 (23 – 33) | 0.47 |
| **Parity, n (%)** |  |  |  | 0.93 |
| 0 | 1,917 (51) | 234 (51) | 1,683 (51) |  |
| 1 | 1,182 (31) | 141 (31) | 1,041 (31) |  |
| 2+ | 694 (18) | 87 (19) | 607 (18) |  |
| Unknown | 629 | 101 | 528 |  |
| **Previous Preterm Deliveries, n (%)** |  |  |  | <0.001 |
| 0 | 3,904 (89) | 407 (75) | 3,497 (91) |  |
| 1 | 397 (9.1) | 102 (19) | 295 (7.7) |  |
| 2+ | 83 (1.9) | 31 (5.7) | 52 (1.4) |  |
| Unknown | 38 | 23 | 15 |  |
| **Previous Abortions (SAB + TAB), n (%)** |  |  |  | 0.68 |
| 0 | 2,293 (58) | 275 (56) | 2,018 (58) |  |
| 1 | 1,161 (29) | 151 (31) | 1,010 (29) |  |
| 2+ | 531 (13) | 64 (13) | 467 (13) |  |
| Unknown | 437 | 73 | 364 |  |
| **MTCL, Median (IQR)** | 36 (33 – 40) | 34 (30 – 39) | 36 (33 – 40) | <0.001 |
| Unknown | 352 | 72 | 280 |  |
| **Gestational Age at Delivery, Median (IQR)** | 39.10 (38.00 – 40.10) | 35.00 (32.30 – 36.30) | 39.30 (38.60 – 40.30) | <0.001 |
| **sCL, n (%)** |  |  |  | <0.001 |
| Normal CL | 3,917 (96) | 428 (87) | 3,489 (97) |  |
| Short CL | 153 (3.8) | 63 (13) | 90 (2.5) |  |
| Unknown | 352 | 72 | 280 |  |

<sup>1</sup> Wilcoxon rank sum test; Pearson's Chi-squared test

### Genetic Correlations between Cervical Length Change and Gestational Duration

Bivariate GCTA analysis of cervical change coefficients and gestational duration suggest that genetic factors significantly contribute to the observed phenotypic correlations between these traits. The observed correlations between the intercept, linear, and nonlinear change coefficients (*r*_Intercept-Linear_ = 0.850 [95% CI 0.846, 0.854], *r*_Intercept-Nonlinear_ = -0.658 [95% CI -0.670, -0.646], *r*_Linear-Nonlinear_ = -0.642 [95% CI -0.654, -0.629]) indicated that a shorter midtrimester cervical length (i.e., intercept) was associated with a more rapid rate of cervical shortening that begins earlier in pregnancy (**Figure 3**). There was significant overlap between the genetic factors influencing cervical change coefficients and gestational age at delivery (*r_g_*, _Intercept-GAD_ = 0.714 [95% CI 0.419, 0.873]; *r_g_*, _Linear-GAD_= 0.982 [95% CI 0.951, 0.993]; *r_g_*_, Nonlinear-GAD_ = -0.490 [95% CI -0.766, -0.062] (**Figure 3, Table S2**). Moreover, the genetic correlation between measures of gestational duration, GAD and sPTB (*r_g_*, _GAD-sPTB_ = -0.600 [95% CI -1.114, -0.086]), was similar to another reported estimate (Solé-Navais *et al*., 2023). These results confirm a genetic correlation between changes in cervical length and gestational age at delivery, suggesting they share underlying biological mechanisms.

### Genome-Wide Associations with Gestational Duration

An exploratory genome-wide analysis was conducted to identify potential genetic mechanisms associated with gestational duration. Two variants were found to be significantly associated with GAD using a genome-wide significance threshold of 1.6×10^−8^ (**Figure 4A**). SNPs rs75296056 and rs185443461 on Chromosome 18 (p = 1.85×10^−9^) are located near the gene encoding the Ras Like Without CAAX 2 (RIT2) protein. RIT2 is a calmodulin-binding GTPase that belongs to the RAS superfamily, and is thought to play a role in cellular signaling, calcium dynamics, dopamine trafficking, and autophagy (Tebar *et al*., 2020; Obergasteiger *et al*., 2023). While the relationship between RIT2 and gestational duration is not clear, it may promote the maintenance of pregnancy by regulating the autophagy-lysosomal pathway, a cellular process of protein degradation that provides energy and nutrients to maintain cellular homeostasis under stressful environmental conditions (Obergasteiger *et al*., 2023). Autophagic activity is essential to the processes of embryonic development, implantation, and placental development in the hypoxic and low-nutrient intrauterine environment of early pregnancy. Dysregulation of placental autophagy has been linked to several pregnancy complications, including miscarriage, gestational hypertension, gestational obesity, gestational diabetes, intrauterine growth restriction, and premature birth (Obergasteiger *et al*., 2023; Zhou *et al*., 2024). In total, 4,410 candidate SNPs, 400 independent lead SNPs, 329 genomic risk loci, and 950 mapped genes were identified using a “suggestive” association threshold of 1×10−5 (**Figure 4A**). The QQ plots for the GWAS and gene-based testing (**Figure S1**) show an acceptable level of genomic inflation (λ_SNP_ = 1.32). These results are consistent with a polygenic architecture, wherein many genes each contribute a small amount to interindividual variation in gestational duration. Genome-wide analysis in this cohort identified two novel loci significantly associated with gestational age at delivery, which were mapped to genes with functions relevant to the maintenance of pregnancy.

**Figure 4.**
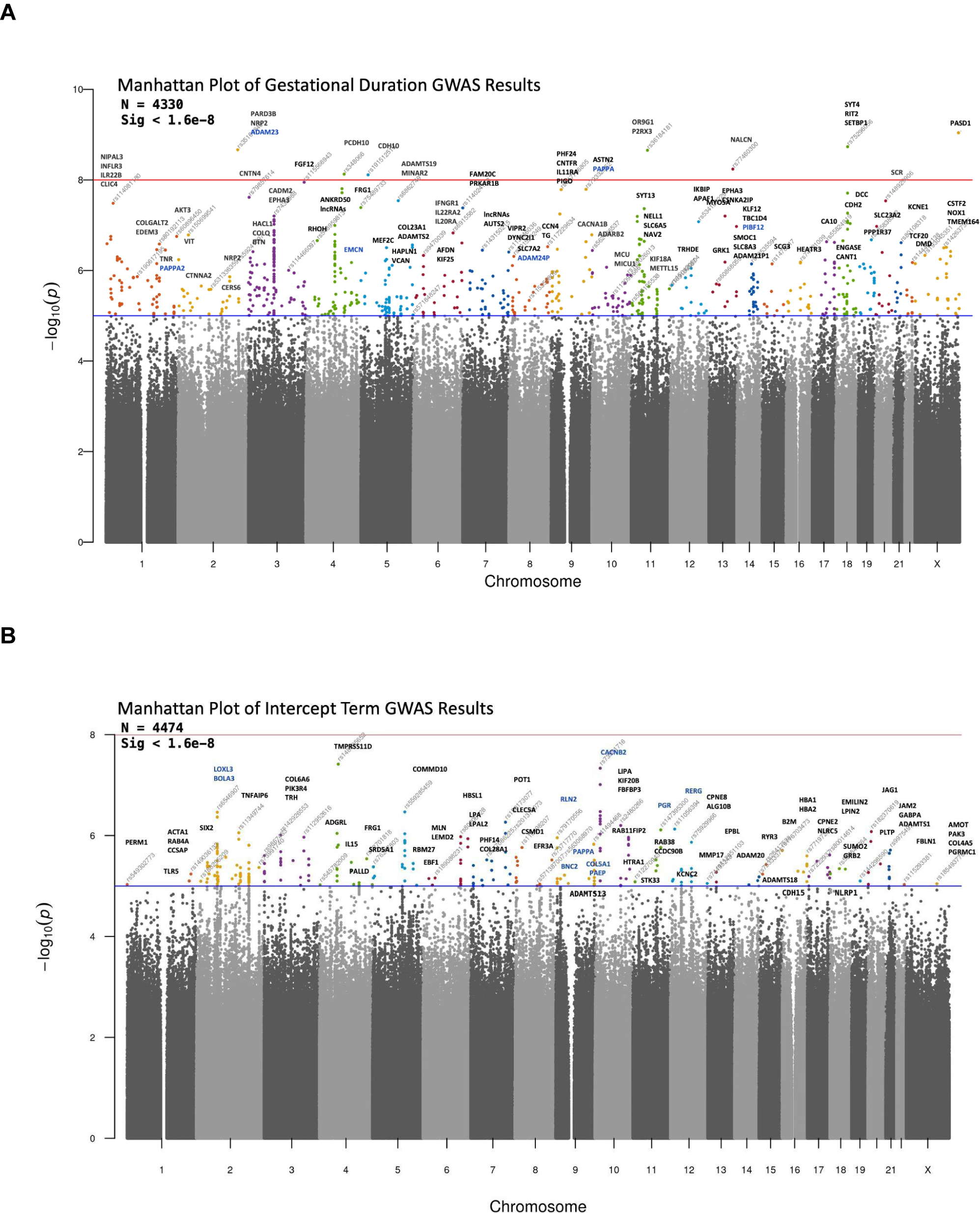
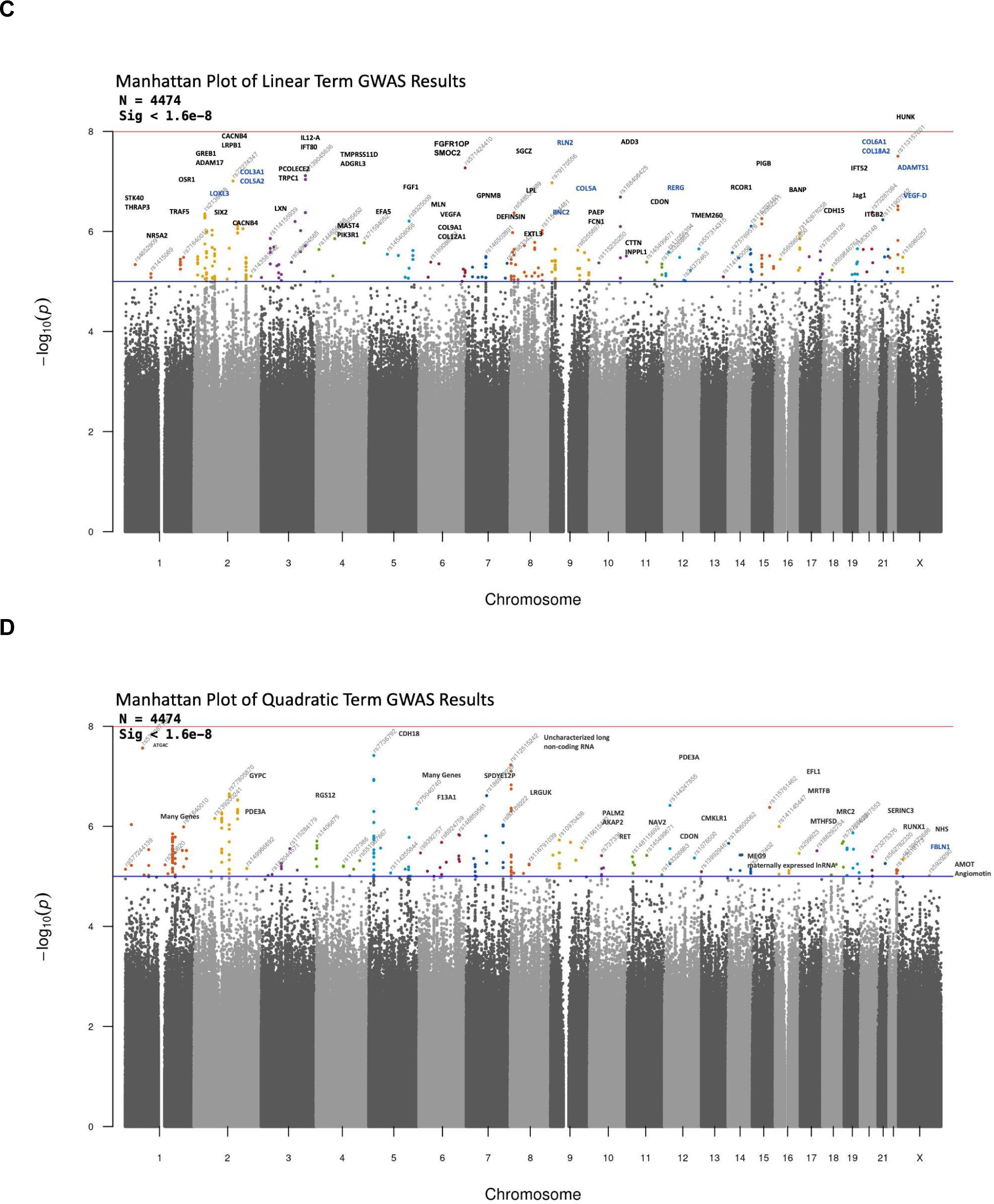
Genome Wide Associations with Cervical Change Coefficients Gestational Outcomes. Manhattan plot (N = 4,423) showing the location and genome-wide association results for SNPs where the red line delineates the genome-wide significance threshold, and the blue line delineates suggestive significance threshold for cervical change coefficients, **A** intercept coefficient (I) **B** linear change coefficient (L), **C** nonlinear change coefficient (Q), and **D** gestational duration (GAD)

### Genome-Wide Associations with Cervical Change

Exploratory genome-wide analyses were also performed to identify genetic associations with measures of cervical change during pregnancy. While no associations met the estimated threshold for genome-wide significance in populations with African ancestry (p < 1.6×10^−8^), associations with two did surpass the standard Bonferroni correction for multiple testing in GWAS (p < 5.0×10^−8^). Using this threshold, rs148405652 on Chromosome 4 (p = 3.85×10^−8^) showed a strong association with estimated mid-trimester cervical length (i.e., intercept coefficient) (**Figure 4B**). This variant maps to the Trypsin-like serine protease gene (TMPRSS11D), a Type II transmembrane serine protease that is involved in regulating many cellular processes relevant to cervical remodeling, including activation of metalloproteases, degradation of extracellular matrix components, fibroblast-to-myofibroblast differentiation, epithelial-mesenchymal transition, microvascular angiogenesis, mobilization of intracellular Ca^2+^, and activation latent growth factors, cytokines and hormones (Antalis *et al*., 2011; Menou *et al*., 2017).

Previously known as human airway trypsin-like protease (HAT), TMPRSS11D/HAT is highly expressed in cervical and vaginal tissue samples and has been detected in the proteome of cervical-vaginal fluid(Uhlén *et al*., 2015; Muytjens *et al*., 2017; GTEx Consortium, 2020). This versatile, membrane-anchored serine protease is linked to a pro-inflammatory immune response and increased mucus production in tissues of the respiratory, lymphoid, and gastrointestinal systems (Menou *et al*., 2017) and has been shown to modulate the pathogenicity, spread, and tissue-specificity of viral infections (Yamaoka *et al*., 1998). TMPRSS11D/HAT may therefore play a crucial role in supporting the adaptive immune and innate barrier functions of mucosal surfaces in the female reproductive tract (Chokki *et al*., 2005; Li *et al*., 2014; Sheng and Hasnain, 2022).

The midtrimester cervical length estimate was also associated with rs73591716 on Chromosome 10 (p = 4.63×10^−8^) (**Figure 4B**). This variant maps to the gene encoding calcium voltage-gated channel auxiliary subunit beta 2 (CACNB2), a subunit of a voltage-dependent calcium channel protein that controls the influx of calcium ions in cardiomyocytes, regulating the rate and contractile force of cardiomyocytes (Papa *et al*., 2022). This gene has been previously associated with Brugada syndrome, an inherited condition that predisposes patients to fatal cardiac arrhythmias (Antzelevitch *et al*., 2007; Watanabe and Minamino, 2016; Moras *et al*., 2023), and risk for severe psychiatric disorders (Andrade *et al*., 2019). Calcium ion (Ca^2+^) signaling plays a fundamental role in regulating uterine myometrial activity during labor and delivery, and calcium channel blockers are used clinically as tocolytic drugs to suppress uterine contractions and delay preterm labor (Gáspár and Hajagos-Tóth, 2013; Flenady *et al*., 2014; Wray *et al*., 2021). Notably, a candidate gene study of in myometrial samples from PPROM patients reported that the expression of several beta subunits of the L-type calcium channel CACNB was significantly higher in patients who delivered preterm compared to full-term deliveries (Kuć *et al*., 2012).

An association was also observed between the linear coefficient and rs139045636 on Chromosome 3 (p = 7.67×10^−8^) which is located within the gene encoding Interleukin 12A + Antisense RNA (*IL-12A-AS1*) and near the gene encoding Interleukin 12A (*IL12A*) **Figure 4C**). Interleukin-12 is a pro-inflammatory cytokine produced primarily by myeloid cells, such as dendritic cells and macrophages, which induces interferon-γ (IFNγ) expression and promotes T helper 1 (Th1) differentiation, among other functions (Trinchieri, 2003; Tait Wojno *et al*., 2019). During pregnancy, there is a shift from the pro-inflammatory microenvironment associated with pregnancy establishment to a homeostatic state that persists for most of gestation to promote maternal-fetal immune tolerance (Gomez-Lopez *et al*., 2022). A breakdown of such maternal-fetal tolerance has therefore been associated with adverse pregnancy outcomes, including preterm labor (Hollier *et al*., 2004; Gomez-Lopez *et al*., 2022). Molecular studies suggest that IL-12 concentrations in cervical secretions are predominantly the result of local cytokine production (Castle *et al*., 2002). Interestingly, cervical concentrations of IL-12 have been associated with many maternal risk factors for a short cervix and preterm labor, including age, parity, genital tract infection, and a high vaginal pH (Hildesheim *et al*., 1999; Crowley-Nowick *et al*., 2000; Gravitt *et al*., 2003).

A threshold of 1×10^−5^ was then used to identify “suggestive” SNPs for exploratory analyses. In total, 150 independent genomic loci containing 300 genes were identified at this threshold across all three cervical length coefficients describing cervical change during pregnancy (**Figures 4B-4D**). Of these, several suggestive SNPs were mapped to genes involved in key hormonal pathways for pregnancy; namely, Low Density Lipoprotein (LDL) Receptor Related Protein 1B (*LRP1B*) (rs141249340, p = 9.81×10^−8^), Relaxin 2 (*RLN2*) (rs35460272, p = 1.06×10^−7^), RAS like estrogen regulated growth inhibitor (RERG) (s11056394, p = 7.45×10^−7^), the Progesterone Receptor (*PGR*) (rs147395300, p = 7.73×10^−7^), Growth Regulating Estrogen Receptor Binding 1 (*GREB1*) (rs13431344, p = 3.64×10^−6^), and pregnancy-associated plasma protein A (*PAPPA*) (rs7865534, p = 7.14×10^−6^). Suggestive associations were also identified between cervical change coefficients and several proteins involved in connective tissue organization, including SPARC-Related Modular Calcium-Binding Protein 2 (SMOC2)(rs571424410, p = 5.36×10^−8^), ADAM metallopeptidase with thrombospondin type 1(ADAMTS1)(rs113157601, p = 3.14×10^−8^), Lysyl Oxidase Like 3 (*LOXL3*) (rs6546907, p = 3.46×10^−7^), Matrix Metallopeptidase 17 (*MMP17*) (rs74495243, p = 8.92×10^−6^), Fibulin 1 (*FBLN1*) (rs115293381, p = 9.37×10^−6^), and fibrillar collagens *COL3A1* and *COL5A2* (rs9776810, p = 9.74×10^−6^).

### Functional Annotation and Gene-Set Enrichment Results

Functional annotations and enrichment analyses were performed on the variants associated with cervical change. Genes mapped to suggestive hits showed downregulated expression in GTEx v8 endocervical tissue (**Figure S2**). Gene-set enrichment profiling of all genes mapped to suggestive SNPs for cervical change resulted in significant enrichment of molecular and functional domains based on a false discovery rate (FDR) of 5% **(Tables S3-S11**). Identified Kegg and Gene Ontology (GO) pathways corresponded to biological processes and molecular functions related to growth factors, energy metabolism, hormonal signaling, immune response, and inflammation (**Tables S7-S10**), including the Interleukin-2 pathways (p = 6.56×10^−6^) and the Interleukin-20 (p = 9.12×10^−6^), Interferon (p = 1.97×10^−5^), and Vascular Endothelial Growth Factor (*VEGFA*, p = 4.88×10^−4^) signaling pathways. Glucose and lipid metabolism (p = 1.57×10^−4^) are vital for fetal development and energy homeostasis during pregnancy, and alterations in these metabolic pathways could negatively impact fetal growth and development, affecting gestational length (Parrettini *et al*., 2020; Jovandaric *et al*., 2023). Estrogen signaling (p = 1.91×10^−4^) plays a significant role in preparing the body for labor and delivery by promoting the production of prostaglandins, lipid compounds that mediate cervical ripening and stimulate myometrial contractions to initiate labor (Socha *et al*., 2022). Likewise, activation of immune cells and the release of cytokines contributes to an inflammatory response in the uterus that is conducive to the onset of labor (Yockey and Iwasaki, 2018).

Differentially expressed gene sets from 54 tissue types (GTEx v8) suggested that many of the mapped genes are differentially expressed in endocervical cells (**Figure S2**). In contrast to the ectocervix, which protrudes into the vagina and is covered by stratified squamous epithelial cells, the endocervix is composed of the columnar epithelial cells that line the cervical canal and internal os and may be influenced by molecular and hormonal signaling from the chorioamniotic membranes (Mendelson *et al*., 2017; Yellon, 2019; Socha *et al*., 2022). Although these results suggest that many of the mapped genes could be expressed in endocervical tissue, the differential regulation results should be interpreted with caution as the 10 cervical tissue samples included in GTEx V8 were collected from non-pregnant donors, many of whom were likely postmenopausal, and thus do not reflect gene expression changes that occur during pregnancy.

## DISCUSSION

Proper cervical length remodeling throughout pregnancy is integral to successfully maintaining a pregnancy and preparing for delivery. While there are many suspected factors that can precipitate early labor (Romero *et al*., 2014; Strauss *et al*., 2018), pathological shortening and effacement of the cervix during pregnancy provides a clear mechanism of preterm birth (Iams *et al*., 1996; Myers *et al*., 2015). This study was designed to provide the first assessment of the genetic architecture of cervical length and quantify its genetic relationship to gestational duration. Low-pass sequencing experiments allowed for the estimation of the number, effect size, and population frequency of genetic variants essential for heritability studies of cervical length and its rate of change across pregnancy.

The findings of the current study have contributed two fundamental insights. First, genetic factors accounted for approximately 50% of inter-individual differences in cervical change during pregnancy. These heritability estimates are consistent with a highly polygenic, complex trait influenced by both genetic and environmental factors (Romero *et al*., 2014; Strauss *et al*., 2018). The estimated heritability of gestational duration outcomes and other biometric measurements were consistent with published estimates for these traits obtained by twin and family and SNP-based estimates in other studies (York *et al*., 2014; Zhang *et al*., 2017; Guo *et al*., 2021; Yengo *et al*., 2022). Specifically, the estimated SNP-based maternal heritability of GAD from this study of self-identified African American women (*h^2^* = 15.8%) was similar to the only estimate available from an African American twin and family cohort (*h^2^* = 13.8%) (York *et al*., 2010). The tendency for SNP-based heritability estimates to be lower than their twin-based counterparts (Friedman *et al*., 2021) was not observed and preliminarily suggests that common genetic alleles, in contrast to rare or structural variants, accounted for the majority of genetic variation in gestational age at birth.

Second, the robust genetic correlations estimated among cervical change and gestational duration traits suggested that a large proportion of the genetic factors contributing to cervical change also influenced gestational duration. Estimates of the genetic correlation along with the trait-specific heritability can be used to provide an approximation of the observed bivariate trait correlations expected to be due to correlated genetic factors (Plomin *et al*., 2012). For example, the observed trait correlation between GAD and the MTCL (i.e., intercept) measures was 0.232 of which 87.1% was estimated to be due to shared genetic factors. Similarly, the GAD-linear (*r*_GAD-Linear_ = 0.246) and GAD-nonlinear (*r*_GAD-Nonlinear_ = -0.148) trait correlations were estimated to be almost entirely due to genetic factors, near 100% and 93.9%, respectively. A meta-analysis of genetic correlations across human diseases and traits confirms that a genetic correlation (*r_g_*) of 0.30 or higher is a reasonable expectation for two traits with a robust epidemiological association, such as cervical length and gestational duration (Bulik-Sullivan *et al*., 2015). It is worth noting that the replication of the GAD-sPTB genetic correlation in the current study (*r_g_* = -0.600 [95% CI -1.114, -0.086]), which was first described by Solé-Navais *et. al.* (*r_g_* = -0.62 [95% CI -0.72, -0.51]) (Solé-Navais *et al*., 2023), does not confirm the genetic similarity of these traits but rather indicates the loss of genetic information after dichotomizing GAD (York *et al*., 2013).

The gene set enrichment and pathway analysis of GWAS summary statistics have shed light on biological mechanisms involved in cervical changes during pregnancy and gestational duration. Pathway enrichment analysis of suggestive variants in this cohort revealed an increased burden in molecular processes related to hormone and growth factor signaling, energy metabolism, tissue remodeling, inflammation, and immune regulation. These biological pathways have been previously implicated in the onset of labor in both premature and term birth. Genomic signals near genes involved in insulin, progesterone, and estrogen signaling, and various cytokines and growth factors like IL-2, IL-20, VEGFA, and interferon signaling were also identified. These results align with current knowledge regarding hormonal influences and responses in cervical tissue, and could offer a potential mechanistic explanation for why intervention with vaginal progesterone might help prolong gestation and prevent preterm birth in women with a short cervix.

Several variants were found to be associated with cervical change and gestational duration. RLN2 is an ovarian hormone that promotes loosening of connective tissues in the pelvic ligaments and the cervix to allow for easier passage of the infant during labor and delivery (Parry and Vodstrcil, 2007). Similar to progesterone, relaxin is thought to promote pregnancy maintenance and uterine quiescence to prevent spontaneous abortions and the premature onset of labor (Goldsmith and Weiss, 2009). Although studies addressing the specific functions of RERG and GREB1 in pregnancy are limited, these proteins are thought to play a role in regulating hormone-responsive pathways in reproductive tissues (Finlin *et al*., 2001; Key *et al*., 2006; Deschênes *et al*., 2007; Mohammed *et al*., 2013). Other hormonally-responsive genes identified herein include LoxL3, which is shown to be physiologically regulated by estrogen in mice (Ozasa *et al*., 1981; Akins *et al*., 2011; Mahendroo, 2012), and FBLN1, a calcium-binding secreted glycoprotein with repeated epidermal-growth-factor-like domains that is also regulated by estrogen (Argraves *et al*., 1990; Clinton *et al*., 1996; Moll *et al*., 2002).

A number of other genes identified herein are implicated in tissue remodeling processes in the context of pregnancy, including MMP17, a membrane-type matrix metalloproteinase that degrades components of the extracellular matrix during tissue remodeling related to embryonic development and reproduction (Yip *et al*., 2019). Fibulins are associated with the formation and rebuilding of extracellular matrices and elastic fibers, including connective tissue disorders in humans (de Vega *et al*., 2009), and FBLN1 has been associated with HPV infection leading to cervical precancerous lesions and cervical carcinoma (Hao *et al*., 2021). COL3A1 and COL5A2 produce alpha chains for a low abundance fibrillar collagens, and mutations in these gene are associated with Ehlers-Danlos syndrome, types I and II, which themselves are associated with an increased risk for cervical insufficiency, sPTB, and PPROM (Modi *et al*., 2017; Strauss *et al*., 2018). While its specific function during pregnancy is not understood, LRP1B has been implicated in modulating inflammatory responses and and may interact with extracellular matrix proteins or cell surface receptors to regulate cell adhesion and migration in the uterus and placenta during pregnancy (Haas *et al*., 2011; Sizova *et al*., 2023). Finally, PAPPA encodes a metalloproteinase that regulates the Insulin-like Growth Factor (IGF) pathway (Oxvig, 2015), and low serum levels of PAPPA in the first trimester have been associated with an increased risk of many adverse pregnancy outcomes, including preterm birth, low birth weight, preeclampsia, miscarriage, and stillbirth (Smith *et al*., 2002; Kirkegaard *et al*., 2010).

The current study was designed to characterize several features of the genetic architecture of cervical change during pregnancy, but was not designed to identify a comprehensive set of common SNP effects. Larger sample sizes will be needed to detect robust statistical associations for individual genetic variants. This study did not address the influence of fetal genes, the vaginal microbiome, or the broader set of environmental and sociodemographic factors that may also contribute to cervical change during pregnancy. Additionally, further study is required to understand the mechanisms behind the generation of genetic correlations between these traits, which could be due to different forms of pleiotropy, linkage disequilibrium, or uncontrolled sources of bias (Solovieff *et al*., 2013).

Previous genetic studies of preterm birth have focused on individuals of European ancestry in Scandinavia and the Americas, limiting the applicability of their findings across diverse global populations. This project is the first large genomic study of birth outcomes in a cohort of US-based women with African ancestry, who are underrepresented in genomic studies, and suffer disproportionately higher rates of sCL and sPTB. Conducting research in a high-risk and underrepresented population could contribute to more effective, personalized prevention strategies for patients from a similar genetic and sociodemographic background.

Collectively, the data herein provide evidence for a substantial genetic influence on changes in cervical length throughout pregnancy. Moreover, such genetic factors are highly correlated with those influencing pregnancy duration, implying shared causal loci between these traits. By undertaking functional and pathway annotation of the observed genetic variants, we identified multiple individual genes and signaling pathways that may be implicated as determinants of cervical changes and gestational duration. Although genetic correlations have not been widely applied to birth outcomes research, they have been extensively used in the fields of psychiatric genetics and endocrinology/cardiology to predict genetic liability for correlated outcomes, enhance causal and mediation analyses, and increase statistical power for multi-trait genetic association studies(Bulik-Sullivan *et al*., 2015). Thus, establishing the shared genetic risk between cervical shortening and gestational duration may improve the utility of cervical length as a screening tool, provide targets for future pharmacogenetic studies, and encourage broader screening programs to identify women with a short cervix who would benefit from clinical interventions, with the ultimate goal of preventing preterm birth.

## DATA AVAILABILITY

All data produced in the present study are available upon reasonable request to the authors.

## AUTHOR’S ROLES

T.P.Y., J.F.S, and R.R. conceived and designed the study. H.M.W, T.P.Y., A.L.T., S.J.L., and B.T.W. contributed to analysis of the data. R.R., S.S.H., T.C., S.B., N.G.L., and P.C. acquired the data and biospecimens. T.P.Y. and H.M.W. developed the manuscript draft. J.F.S, B.T.W., A.L.T., N.G.L., and R.R. critically revised the article. All authors gave final approval of the version to be published.

## FUNDING

This research was supported by the Perinatology Research Branch, Division of Obstetrics and Maternal-Fetal Medicine, Division of Intramural Research, *Eunice Kennedy Shriver* National Institute of Child Health and Human Development, National Institutes of Health, United States Department of Health and Human Services (NICHD/NIH/DHHS) (Contract No. HHSN275201300006C). RR has contributed to this work as part of his official duties as an employee of the United States Federal Government. A.L.T., N.G.L., and S.S.H. were also supported by the Wayne State University Perinatal Initiative in Maternal, Perinatal and Child Health. Additional support was provided from the March of Dimes Prematurity Research Center at the University of Pennsylvania (22-FY18-812).

## CONFLICT OF INTEREST

The funders had no influence on the data collection, analyses or conclusions of the study. There is no conflict of interest to declare.

## SUPPLEMENTAL MATERIALS

**Table S1.** Heritability estimates for cervical length trajectory coefficients, and gestational duration outcomes, height, and BMI. V_G_ represents a trait’s total genetic variance; V_E_ represents a trait’s total environmental variance; V_P_ represent a trait’s total phenotypic variance; h^2^ (V_G_ / V_p_) represents a trait’s estimated heritability, or the proportion of phenotypic variance explained by all SNPs for each trait. Literature values of SNP-based heritability are included for comparison for traits that have been studied in other cohorts. (*I*: intercept; *L*: linear term; *Q*: quadratic term).

| Trait | $V_G$ | $V_G$ SE | $V_E$ | $V_E$ SE | $V_P$ | $V_P$ SE | $h^2$ | $h^2$ SE | Literature $h^2$<br>(SNP-based GCTA) | Literature SE |
| --- | --- | --- | --- | --- | --- | --- | --- | --- | --- | --- |
| **I** | 9.581 | 1.797 | 9.242 | 1.737 | 18.823 | 0.407 | 0.509 | 0.093 | NA | NA |
| **L** | 0.945 | 0.180 | 0.912 | 0.174 | 1.857 | 0.040 | 0.509 | 0.095 | NA | NA |
| **Q** | 0.015 | 0.003 | 0.015 | 0.003 | 0.029 | 0.001 | 0.507 | 0.093 | NA | NA |
| **MTCL** | 16.401 | 6.232 | 33.765 | 6.147 | 50.166 | 1.264 | 0.327 | 0.123 | NA | NA |
| **GAD** | 1.418 | 0.829 | 7.553 | 0.830 | 8.971 | 0.187 | 0.158 | 0.092 | 0.17<br>PMID: 28877031<br>Zhang, 2017 | 0.05 |
| **PTB** | 0.013 | 0.008 | 0.073 | 0.008 | 0.086 | 0.002 | 0.154 | 0.091 | 0.23<br>PMID: 28877031<br>Zhang, 2017 | 0.08 |
| **Height** | 0.004 | 0.000 | 0.002 | 0.000 | 0.005 | 0.000 | 0.675 | 0.086 | 0.46<br>Yengo, 2022<br>PMID: 36224396 | 0.04 |
| **BMI** | 32.918 | 5.292 | 24.228 | 5.075 | 57.146 | 1.203 | 0.576 | 0.090 | 0.22<br>Guo, 2021 | 0.03 |

**Table S2.** Estimates for generic correlations between cervical length growth parameters and gestational duration. Note: trait 1 represents the first trait listed in the trait name (for the correlation between I & GAD, I is trait 1 and GAD is trait 2). Cor_P_ represents the correlation between the phenotypic values of the two traits. V_G_ t1 and V_G_ t2 represent the variation from additive genetic effects for traits 1 and 2, respectively; C_G_t12 represents the genetic covariance between traits 1 and 2; V_E_ t1 and V_E_ t2 represent the residual/environmental variance for traits 1 and 2, respectively; C_E_t12 represents the residual/environmental covariance between traits 1 and 2; V_P_ t1 and V_P_ t2 represent the total phenotypic variation traits 1 and 2, respectively; h^2^ (V_G_ / V_p_) represents the proportion of phenotypic variance explained by all SNPs for each trait; and r_G_ represents the genetic correlation between trait 1 and 2.

| Trait | $Cor_P$ | $Cor_P$ SE | $V_G$ t1 | $V_G$ t2 | $C_G$t12 | $V_E$ t1 | $V_E$ t2 | $C_E$t12 | $V_P$ t1 | $V_P$ t2 | $h^2$ t1 | $h^2$ t1 SE | $h^2$ t2 | $h^2$ t2 SE | $r_G$ | $r_G$ SE |
| --- | --- | --- | --- | --- | --- | --- | --- | --- | --- | --- | --- | --- | --- | --- | --- | --- |
| I & GAD | 0.232 | 0.015 | 10.018 | 1.492 | 2.759 | 9.012 | 7.480 | 0.663 | 19.030 | 8.972 | 0.526 | 0.093 | 0.166 | 0.092 | 0.714 | 0.229 |
| L & GAD | 0.246 | 0.015 | 1.014 | 1.544 | 1.229 | 0.866 | 7.429 | -0.090 | 1.880 | 8.973 | 0.539 | 0.094 | 0.172 | 0.091 | 0.982 | 0.257 |
| Q & GAD | -0.148 | 0.015 | 0.015 | 1.402 | -0.071 | 0.014 | 7.568 | -0.013 | 0.030 | 8.971 | 0.511 | 0.093 | 0.156 | 0.092 | -0.490 | 0.242 |
| I & sPTB | -0.179 | 0.015 | 9.861 | 0.013 | -0.266 | 9.065 | 0.073 | 0.008 | 18.926 | 0.086 | 0.521 | 0.093 | 0.152 | 0.091 | -0.741 | 0.268 |
| L & sPTB | -0.214 | 0.015 | 0.989 | 0.013 | -0.101 | 0.882 | 0.073 | 0.006 | 1.871 | 0.086 | 0.529 | 0.094 | 0.153 | 0.090 | -0.888 | 0.284 |
| Q & sPTB | 0.115 | 0.015 | 0.015 | 0.013 | 0.006 | 0.015 | 0.073 | 0.000 | 0.030 | 0.086 | 0.507 | 0.093 | 0.148 | 0.091 | 0.467 | 0.258 |
| GAD & sPTB | -0.627 | 0.011 | 1.419 | 0.013 | -0.082 | 7.552 | 0.074 | -0.475 | 8.971 | 0.087 | 0.158 | 0.092 | 0.150 | 0.091 | -0.600 | 0.262 |
| I & L | 0.850 | 0.008 | 9.856 | 0.944 | 3.040 | 8.976 | 0.913 | 1.961 | 18.832 | 1.857 | 0.523 | 0.093 | 0.508 | 0.095 | 0.997 | 0.026 |
| I & Q | -0.658 | 0.011 | 9.638 | 0.015 | -0.326 | 9.186 | 0.014 | -0.163 | 18.825 | 0.029 | 0.512 | 0.093 | 0.511 | 0.093 | -0.855 | 0.062 |
| L & Q | -0.642 | 0.011 | 0.934 | 0.015 | -0.106 | 0.922 | 0.014 | -0.044 | 1.856 | 0.030 | 0.503 | 0.094 | 0.512 | 0.092 | -0.891 | 0.065 |

**Table S3.**
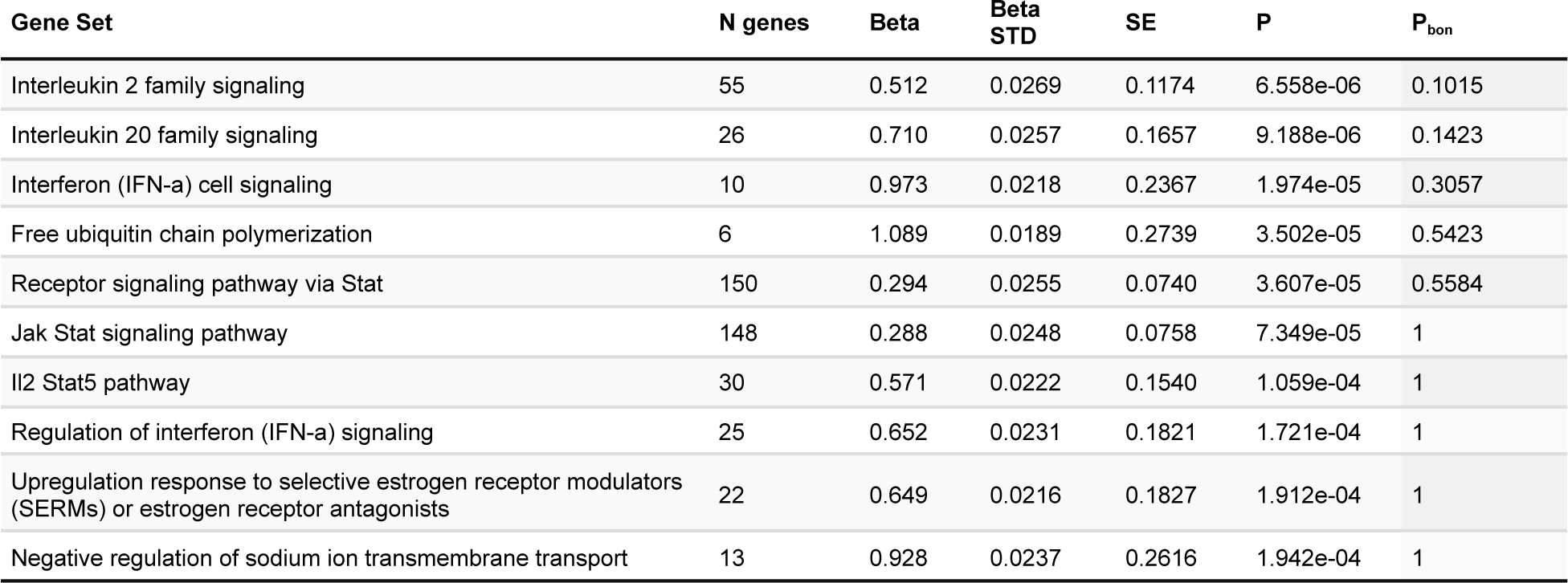
Top 10 gene-set enrichment categories identified using gene-based tests of GWAS summary statistics for the intercept coefficient (I) computed by MAGMA.

| Gene Set | N genes | Beta | Beta STD | SE | P | $P_{bon}$ |
| --- | --- | --- | --- | --- | --- | --- |
| Interleukin 2 family signaling | 55 | 0.512 | 0.0269 | 0.1174 | 6.558e-06 | 0.1015 |
| Interleukin 20 family signaling | 26 | 0.710 | 0.0257 | 0.1657 | 9.188e-06 | 0.1423 |
| Interferon (IFN-a) cell signaling | 10 | 0.973 | 0.0218 | 0.2367 | 1.974e-05 | 0.3057 |
| Free ubiquitin chain polymerization | 6 | 1.089 | 0.0189 | 0.2739 | 3.502e-05 | 0.5423 |
| Receptor signaling pathway via Stat | 150 | 0.294 | 0.0255 | 0.0740 | 3.607e-05 | 0.5584 |
| Jak Stat signaling pathway | 148 | 0.288 | 0.0248 | 0.0758 | 7.349e-05 | 1 |
| Il2 Stat5 pathway | 30 | 0.571 | 0.0222 | 0.1540 | 1.059e-04 | 1 |
| Regulation of interferon (IFN-a) signaling | 25 | 0.652 | 0.0231 | 0.1821 | 1.721e-04 | 1 |
| Upregulation response to selective estrogen receptor modulators (SERMs) or estrogen receptor antagonists | 22 | 0.649 | 0.0216 | 0.1827 | 1.912e-04 | 1 |
| Negative regulation of sodium ion transmembrane transport | 13 | 0.928 | 0.0237 | 0.2616 | 1.942e-04 | 1 |

**Table S4.**
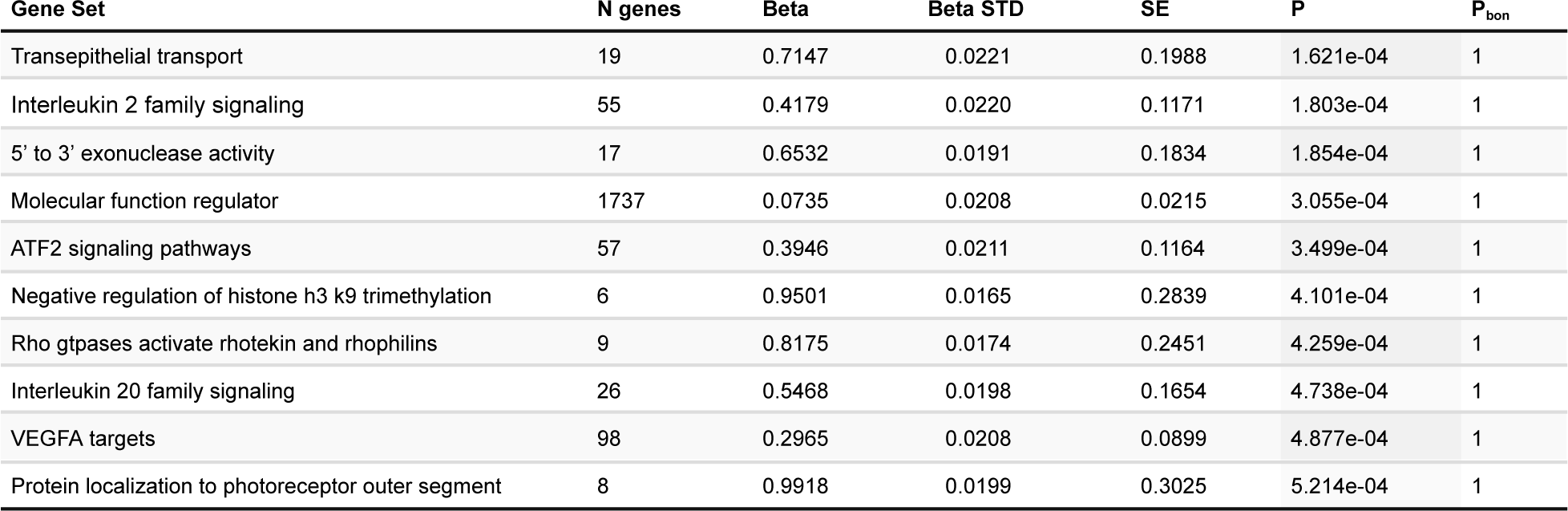
Top 10 gene-set enrichment categories identified using gene-based tests of GWAS summary statistics for the linear change coefficient (L), computed by MAGMA.

**Table S5.** Top 10 gene-set enrichment categories identified using gene-based tests of GWAS summary statistics for the nonlinear change coefficient (Q), computed by MAGMA.

| Gene Set | N genes | Beta | Beta STD | SE | P | P<sub>bon</sub> |
| --- | --- | --- | --- | --- | --- | --- |
| E-box enhancer binding | 49 | 0.4677 | 0.0232 | 0.1225 | 6.773e-05 | 1 |
| Positive regulation of integrin activation | 8 | 1.1064 | 0.0222 | 0.3059 | 1.496e-04 | 1 |
| Lipid phosphorylation | 62 | 0.3824 | 0.0213 | 0.1061 | 1.572e-04 | 1 |
| Interleukin 20 family signaling | 26 | 0.5879 | 0.0213 | 0.1654 | 1.903e-04 | 1 |
| Forebrain radial glial cell differentiation | 7 | 1.1321 | 0.0213 | 0.3273 | 2.720e-04 | 1 |
| Cell migration in hindbrain | 15 | 0.7572 | 0.0208 | 0.2191 | 2.745e-04 | 1 |
| Long chain fatty acid import into cell | 15 | 0.5946 | 0.0163 | 0.1741 | 3.199e-04 | 1 |
| Receptor complex | 389 | 0.1527 | 0.0212 | 0.0451 | 3.563e-04 | 1 |
| Insulin signaling pathway | 136 | 0.2495 | 0.0206 | 0.0738 | 3.600e-04 | 1 |
| Down-regulated after stimulation with galecin-1 (lectin, LGALS1) vs lipopolysaccharide (LPS). | 438 | 0.1345 | 0.0198 | 0.0403 | 4.256e-04 | 1 |

**Table S6.**
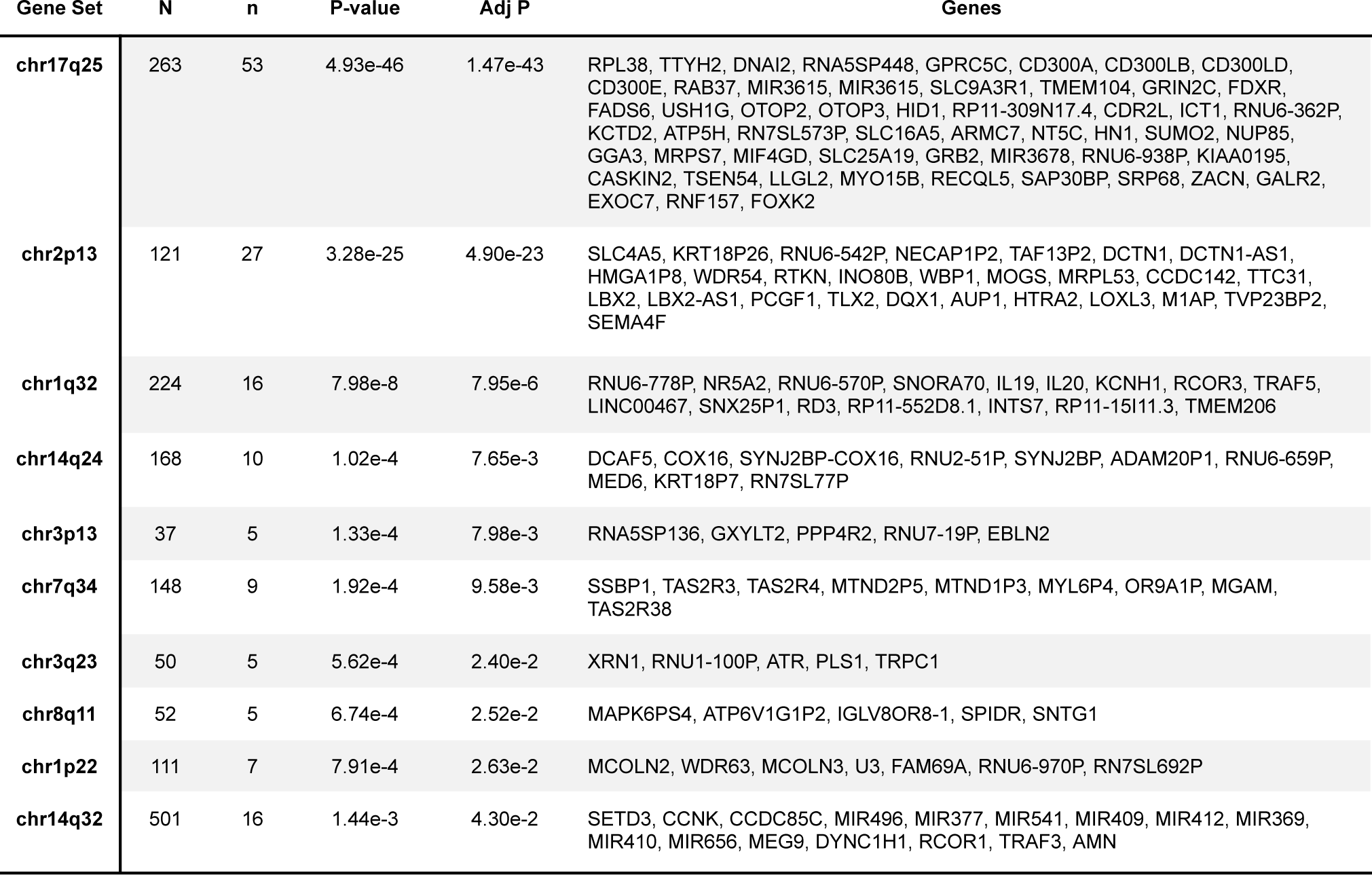
Positional gene enrichment sets, using summary statistics GWA Studies for Cervical Change coefficients (I,L, and Q)

**Table S7.** Kegg Pathway Analysis for Cervical Change coefficients (I,L, and Q)

| Gene Set | N | n | P-value | Adj P | Genes |
| --- | --- | --- | --- | --- | --- |
| Long Term Potentiation | 70 | 6 | 3.69e-4 | 4.42e-2 | GRIN2C, PRKCG, CAMK2D, GRIA2, ITPR3, ADCY8 |
| Taste Transduction | 51 | 5 | 6.16e-4 | 4.42e-2 | ITPR3, TAS2R3, TAS2R4, TAS2R38, ADCY8 |
| Calcium Signaling Pathway | 177 | 9 | 7.13e-4 | 4.42e-2 | GRIN2C, PRKCG, SLC8A1, TNNC2, TRPC1, CAMK2D, ITPR3, ADCY8, GRPR |

**Table S8.** Gene Ontology: Biological Processes for Cervical Change coefficients (I,L, and Q)

| Gene Set | N | n | P-value | Adj P | Genes |
| --- | --- | --- | --- | --- | --- |
| Cellular Macromolecule Localization | 1886 | 50 | 4.64e-6 | 3.30e-2 | ATG4C, WLS, DNM3, RAB4A, NRP1, TUB, DLG2, RAB38, KPNA3, PRKCH, SYNJ2BP, AMN, AP3B2, BANP, RPL38, RAB37, SLC9A3R1, GRIN2C, HID1, NUP85, GGA3, SRP68, CDC37, DNM2, TMED1, CARM1, MARK4, KPTN, NAPA, MYADM, DDX1, COMMD1, RTKN, AUP1, HTRA2, SNAP25-AS1, SRSF6, ACOT8, MRAP, ATR, PLS1, TSPAN5, LEMD2, RIMS1, NUPL2, HIP1, CNTNAP2, MCPH1, SPIDR, APBA1 |
| Regulation Of Cell Morphogenesis | 475 | 20 | 8.97e-6 | 3.30e-2 | DNM3, NRP1, PLXNC1, SLC9A3R1, RNF157, DNM2, MYADM, TLX2, SEMA4F, RTN4R, PARVB, LIMD1, EFNA5, RIMS1, HECW1, DLC1, DOCK5, PTPRD, PALM2, PAK3 |

**Table S9.** Gene Ontology: Molecular Functions for Cervical Change coefficients (I,L, and Q)

| Gene Set | N | n | P-value | Adj P | Genes |
| --- | --- | --- | --- | --- | --- |
| Haptoglobin Binding | 10 | 4 | 6.44e-6 | 1.06e-2 | HBM, HBA2, HBA1, HBQ1 |
| Molecular Carrier Activity | 41 | 6 | 1.76e-5 | 1.45e-2 | KPNA3, HBM, HBA2, HBA1, HBQ1, NUPL2 |
| Oxygen Carrier Activity | 14 | 4 | 2.94e-5 | 1.61e-2 | HBM, HBA2, HBA1, HBQ1 |
| Heparin Binding | 163 | 10 | 7.96e-5 | 3.27e-2 | NRP1, NAV2, BSPH1, ELSPBP1, MSTN, RTN4R, ANXA6, LPA, GPNMB, LPL |
| Oxygen Binding | 36 | 5 | 1.17e-4 | 3.84e-2 | HBM, HBA2, HBA1, HBQ1, ADGB |

**Table S10.**
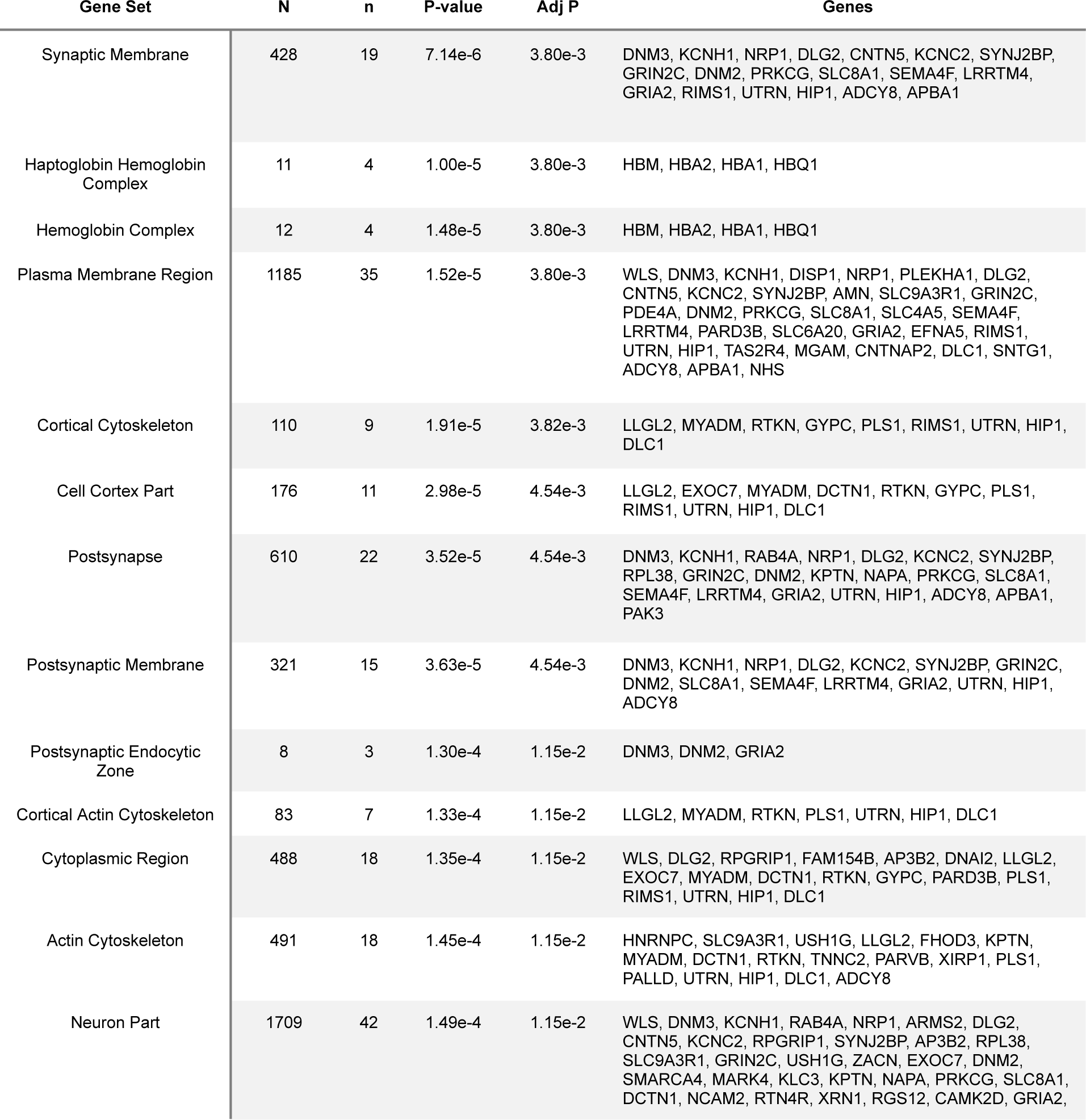
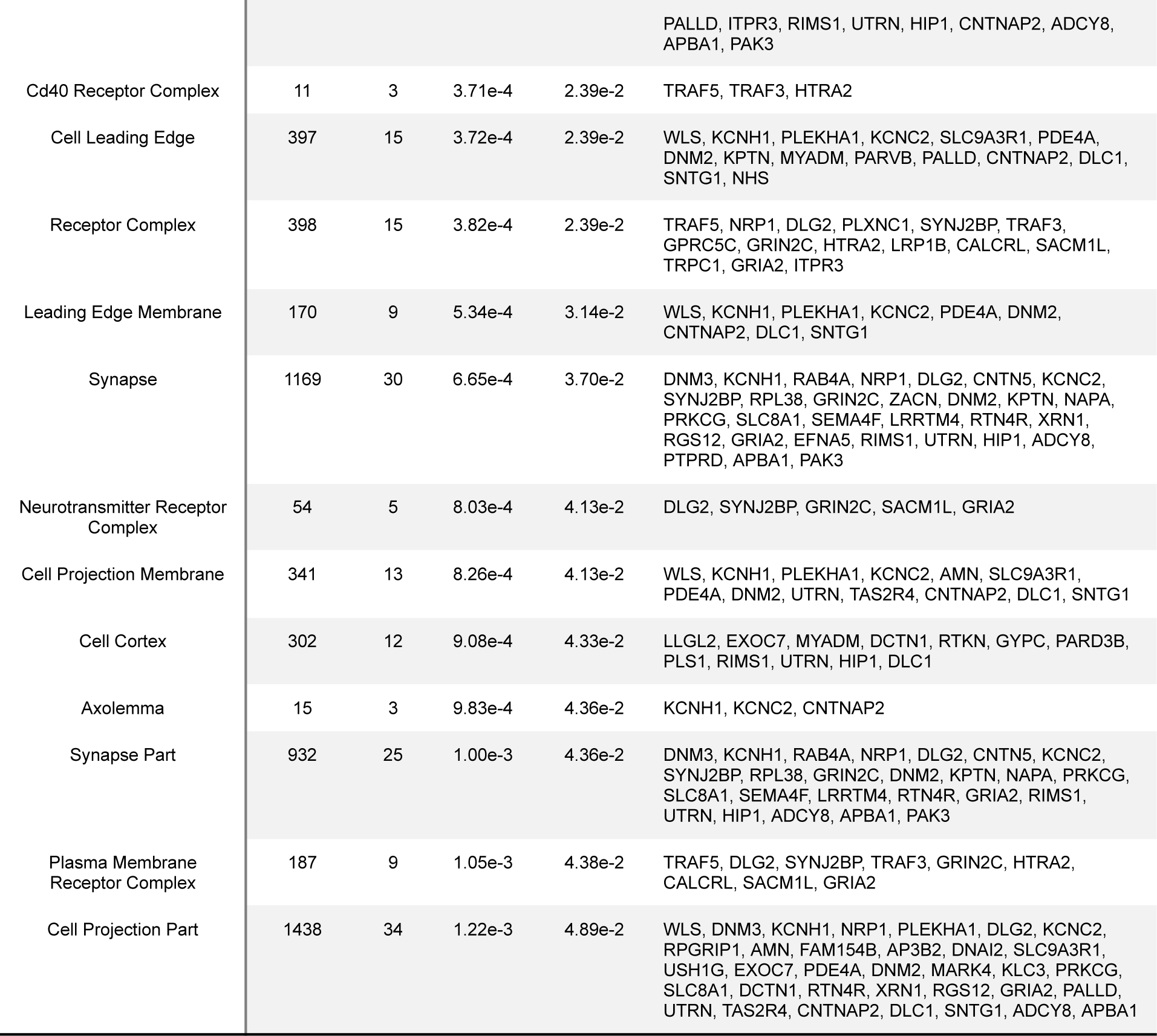
Gene Ontology: Cellular Components for Cervical Change coefficients (I,L, and Q)

| Gene Set | N | n | P-value | Adj P | Genes |
| --- | --- | --- | --- | --- | --- |
| Synaptic Membrane | 428 | 19 | 7.14e-6 | 3.80e-3 | DNM3, KCNH1, NRP1, DLG2, CNTN5, KCNC2, SYNJ2BP, GRIN2C, DNM2, PRKCG, SLC8A1, SEMA4F, LRRTM4, GRIA2, RIMS1, UTRN, HIP1, ADCY8, APBA1 |
| Haptoglobin Hemoglobin Complex | 11 | 4 | 1.00e-5 | 3.80e-3 | HBM, HBA2, HBA1, HBQ1 |
| Hemoglobin Complex | 12 | 4 | 1.48e-5 | 3.80e-3 | HBM, HBA2, HBA1, HBQ1 |
| Plasma Membrane Region | 1185 | 35 | 1.52e-5 | 3.80e-3 | WLS, DNM3, KCNH1, DISP1, NRP1, PLEKHA1, DLG2, CNTN5, KCNC2, SYNJ2BP, AMN, SLC9A3R1, GRIN2C, PDE4A, DNM2, PRKCG, SLC8A1, SLC4A5, SEMA4F, LRRTM4, PARD3B, SLC6A20, GRIA2, EFNA5, RIMS1, UTRN, HIP1, TAS2R4, MGAM, CNTNAP2, DLC1, SNTG1, ADCY8, APBA1, NHS |
| Cortical Cytoskeleton | 110 | 9 | 1.91e-5 | 3.82e-3 | LLGL2, MYADM, RTKN, GYPC, PLS1, RIMS1, UTRN, HIP1, DLC1 |
| Cell Cortex Part | 176 | 11 | 2.98e-5 | 4.54e-3 | LLGL2, EXOC7, MYADM, DCTN1, RTKN, GYPC, PLS1, RIMS1, UTRN, HIP1, DLC1 |
| Postsynapse | 610 | 22 | 3.52e-5 | 4.54e-3 | DNM3, KCNH1, RAB4A, NRP1, DLG2, KCNC2, SYNJ2BP, RPL38, GRIN2C, DNM2, KPTN, NAPA, PRKCG, SLC8A1, SEMA4F, LRRTM4, GRIA2, UTRN, HIP1, ADCY8, APBA1, PAK3 |
| Postsynaptic Membrane | 321 | 15 | 3.63e-5 | 4.54e-3 | DNM3, KCNH1, NRP1, DLG2, KCNC2, SYNJ2BP, GRIN2C, DNM2, SLC8A1, SEMA4F, LRRTM4, GRIA2, UTRN, HIP1, ADCY8 |
| Postsynaptic Endocytic Zone | 8 | 3 | 1.30e-4 | 1.15e-2 | DNM3, DNM2, GRIA2 |
| Cortical Actin Cytoskeleton | 83 | 7 | 1.33e-4 | 1.15e-2 | LLGL2, MYADM, RTKN, PLS1, UTRN, HIP1, DLC1 |
| Cytoplasmic Region | 488 | 18 | 1.35e-4 | 1.15e-2 | WLS, DLG2, RPGRIP1, FAM154B, AP3B2, DNAI2, LLGL2, EXOC7, MYADM, DCTN1, RTKN, GYPC, PARD3B, PLS1, RIMS1, UTRN, HIP1, DLC1 |
| Actin Cytoskeleton | 491 | 18 | 1.45e-4 | 1.15e-2 | HNRNPC, SLC9A3R1, USH1G, LLGL2, FHOD3, KPTN, MYADM, DCTN1, RTKN, TNNC2, PARVB, XIRP1, PLS1, PALLD, UTRN, HIP1, DLC1, ADCY8 |
| Neuron Part | 1709 | 42 | 1.49e-4 | 1.15e-2 | WLS, DNM3, KCNH1, RAB4A, NRP1, ARMS2, DLG2, CNTN5, KCNC2, RPGRIP1, SYNJ2BP, AP3B2, RPL38, SLC9A3R1, GRIN2C, USH1G, ZACN, EXOC7, DNM2, SMARCA4, MARK4, KLC3, KPTN, NAPA, PRKCG, SLC8A1, DCTN1, NCAM2, RTN4R, XRN1, RGS12, CAMK2D, GRIA2, |
|  |  |  |  |  | PALLD, ITPR3, RIMS1, UTRN, HIP1, CNTNAP2, ADCY8, APBA1, PAK3 |
| Cd40 Receptor Complex | 11 | 3 | 3.71e-4 | 2.39e-2 | TRAF5, TRAF3, HTRA2 |
| Cell Leading Edge | 397 | 15 | 3.72e-4 | 2.39e-2 | WLS, KCNH1, PLEKHA1, KCNC2, SLC9A3R1, PDE4A, DNM2, KPTN, MYADM, PARVB, PALLD, CNTNAP2, DLC1, SNTG1, NHS |
| Receptor Complex | 398 | 15 | 3.82e-4 | 2.39e-2 | TRAF5, NRP1, DLG2, PLXNC1, SYNJ2BP, TRAF3, GPRC5C, GRIN2C, HTRA2, LRP1B, CALCRL, SACM1L, TRPC1, GRIA2, ITPR3 |
| Leading Edge Membrane | 170 | 9 | 5.34e-4 | 3.14e-2 | WLS, KCNH1, PLEKHA1, KCNC2, PDE4A, DNM2, CNTNAP2, DLC1, SNTG1 |
| Synapse | 1169 | 30 | 6.65e-4 | 3.70e-2 | DNM3, KCNH1, RAB4A, NRP1, DLG2, CNTN5, KCNC2, SYNJ2BP, RPL38, GRIN2C, ZACN, DNM2, KPTN, NAPA, PRKCG, SLC8A1, SEMA4F, LRRTM4, RTN4R, XRN1, RGS12, GRIA2, EFNA5, RIMS1, UTRN, HIP1, ADCY8, PTPRD, APBA1, PAK3 |
| Neurotransmitter Receptor Complex | 54 | 5 | 8.03e-4 | 4.13e-2 | DLG2, SYNJ2BP, GRIN2C, SACM1L, GRIA2 |
| Cell Projection Membrane | 341 | 13 | 8.26e-4 | 4.13e-2 | WLS, KCNH1, PLEKHA1, KCNC2, AMN, SLC9A3R1, PDE4A, DNM2, UTRN, TAS2R4, CNTNAP2, DLC1, SNTG1 |
| Cell Cortex | 302 | 12 | 9.08e-4 | 4.33e-2 | LLGL2, EXOC7, MYADM, DCTN1, RTKN, GYPC, PARD3B, PLS1, RIMS1, UTRN, HIP1, DLC1 |
| Axolemma | 15 | 3 | 9.83e-4 | 4.36e-2 | KCNH1, KCNC2, CNTNAP2 |
| Synapse Part | 932 | 25 | 1.00e-3 | 4.36e-2 | DNM3, KCNH1, RAB4A, NRP1, DLG2, CNTN5, KCNC2, SYNJ2BP, RPL38, GRIN2C, DNM2, KPTN, NAPA, PRKCG, SLC8A1, SEMA4F, LRRTM4, RTN4R, GRIA2, RIMS1, UTRN, HIP1, ADCY8, APBA1, PAK3 |
| Plasma Membrane Receptor Complex | 187 | 9 | 1.05e-3 | 4.38e-2 | TRAF5, DLG2, SYNJ2BP, TRAF3, GRIN2C, HTRA2, CALCRL, SACM1L, GRIA2 |
| Cell Projection Part | 1438 | 34 | 1.22e-3 | 4.89e-2 | WLS, DNM3, KCNH1, NRP1, PLEKHA1, DLG2, KCNC2, RPGRIP1, AMN, FAM154B, AP3B2, DNAI2, SLC9A3R1, USH1G, EXOC7, PDE4A, DNM2, MARK4, KLC3, PRKCG, SLC8A1, DCTN1, RTN4R, XRN1, RGS12, GRIA2, PALLD, UTRN, TAS2R4, CNTNAP2, DLC1, SNTG1, ADCY8, APBA1 |

**Table S11.**
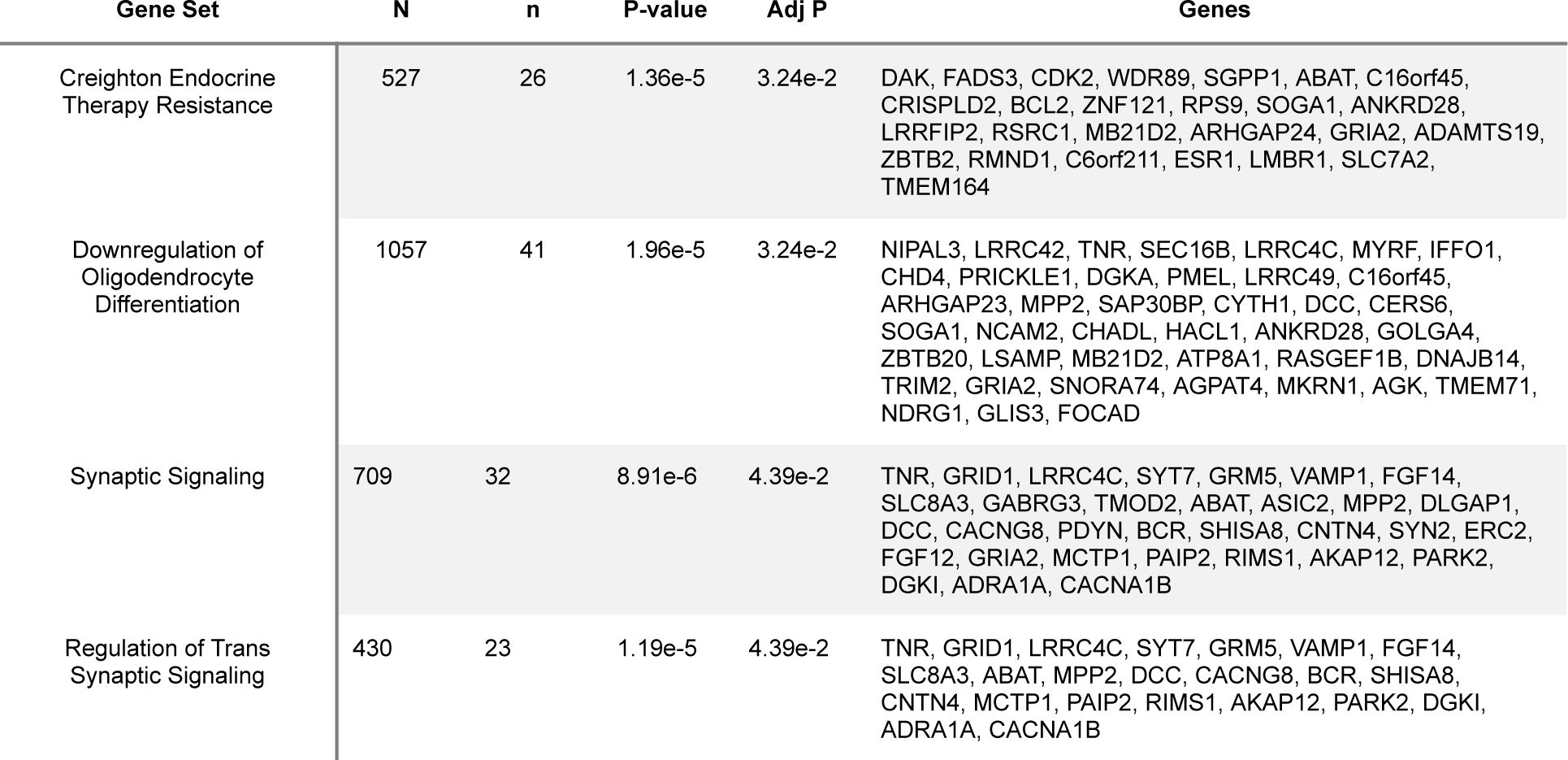
Gene-set enrichment categories identified using gene-based tests of GWAS summary statistics from GWA studies of Cervical Change coefficients (I,L, and Q), computed by MAGMA.

**Table S12.** Gene-set enrichment categories identified using gene-based tests of GWAS summary statistics for GAD, computed by MAGMA.

| Gene Set | N genes | Beta | Beta STD | SE | P | P<sub>bon</sub> |
| --- | --- | --- | --- | --- | --- | --- |
| Neuronal Dense Core Vesicle | 6 | 1.5721 | 0.0273 | 0.3651 | 8.373e-06 | 0.1297 |
| Formation Of Tubulin Folding Intermediates | 26 | 0.5853 | 0.0212 | 0.1449 | 2.683e-05 | 0.4155 |
| Filamin Binding | 13 | 0.8736 | 0.0223 | 0.2316 | 8.120e-05 | 1 |
| Fu Interact With Alkbh8 | 13 | 0.7143 | 0.0183 | 0.1899 | 8.473e-05 | 1 |
| Positive Regulation Of Telomerase Rna Localization To Cajal Body | 15 | 0.6390 | 0.0176 | 0.1797 | 1.888e-04 | 1 |
| Secondary Metabolite Biosynthetic Process | 27 | 0.5598 | 0.0206 | 0.1602 | 2.379e-04 | 1 |
| Upregulation: Aging | 5 | 1.4413 | 0.0229 | 0.4139 | 2.495e-04 | 1 |
| Transport Of Connexons to the Plasma Membrane | 20 | 0.6437 | 0.0204 | 0.1850 | 2.513e-04 | 1 |
| D4-GDI (GDP dissociation inhibitor) Pathway | 12 | 0.9218 | 0.0227 | 0.2705 | 3.274e-04 | 1 |
| H3k4me3 And H3k27me3 | 131 | 0.2536 | 0.0205 | 0.0748 | 3.499e-04 | 1 |

## SUPPLEMENTAL FIGURES

**Figure S1.**
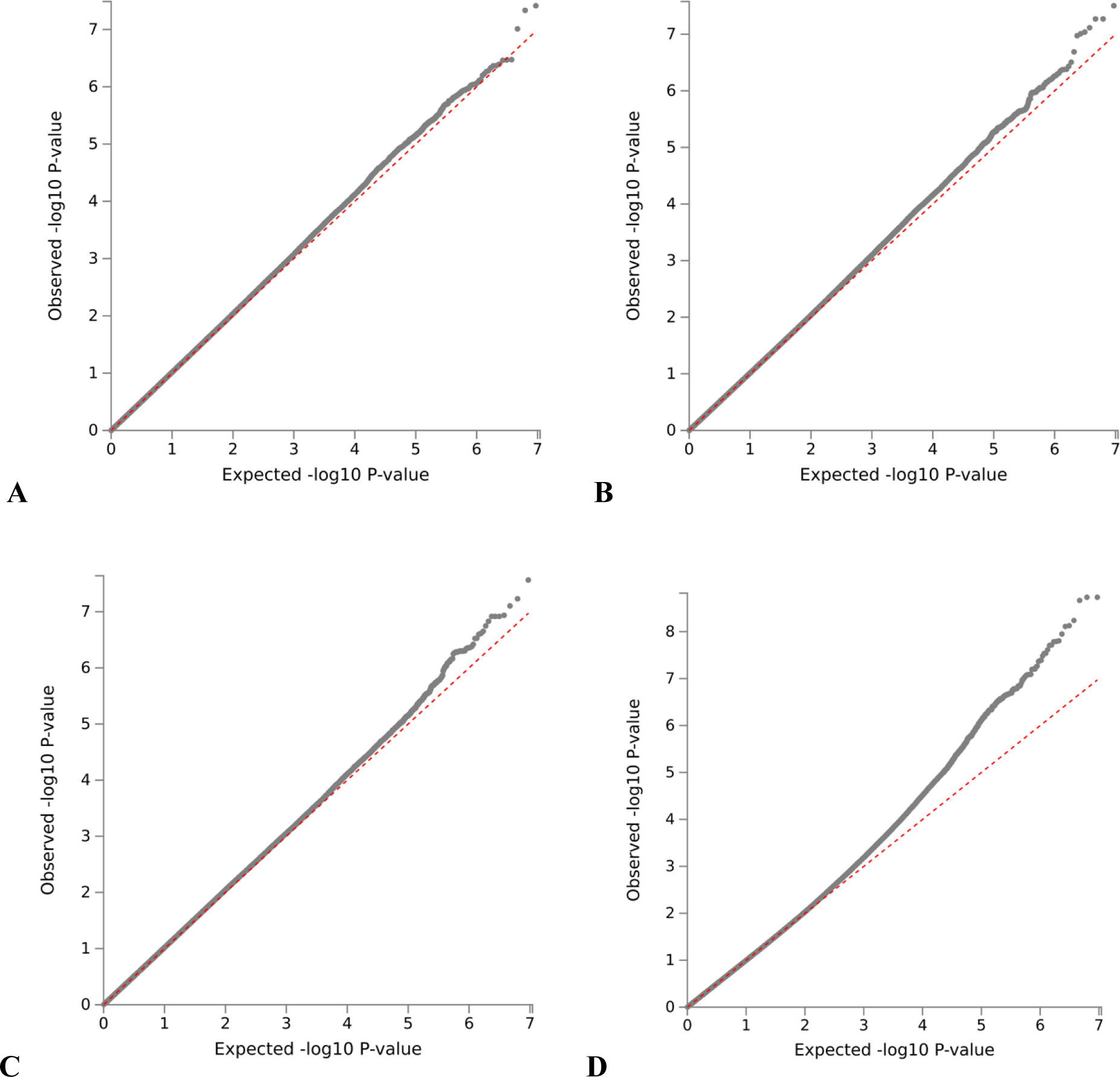
QQ-plots for GWA Studies. **A** QQ-plot for Intercept coefficient (I), **B** QQ-plot for Linear change coefficient (L), **C** QQ-plot for Nonlinear change coefficient (Q), **D** QQ-plot for GAD.

**Figure S2.**
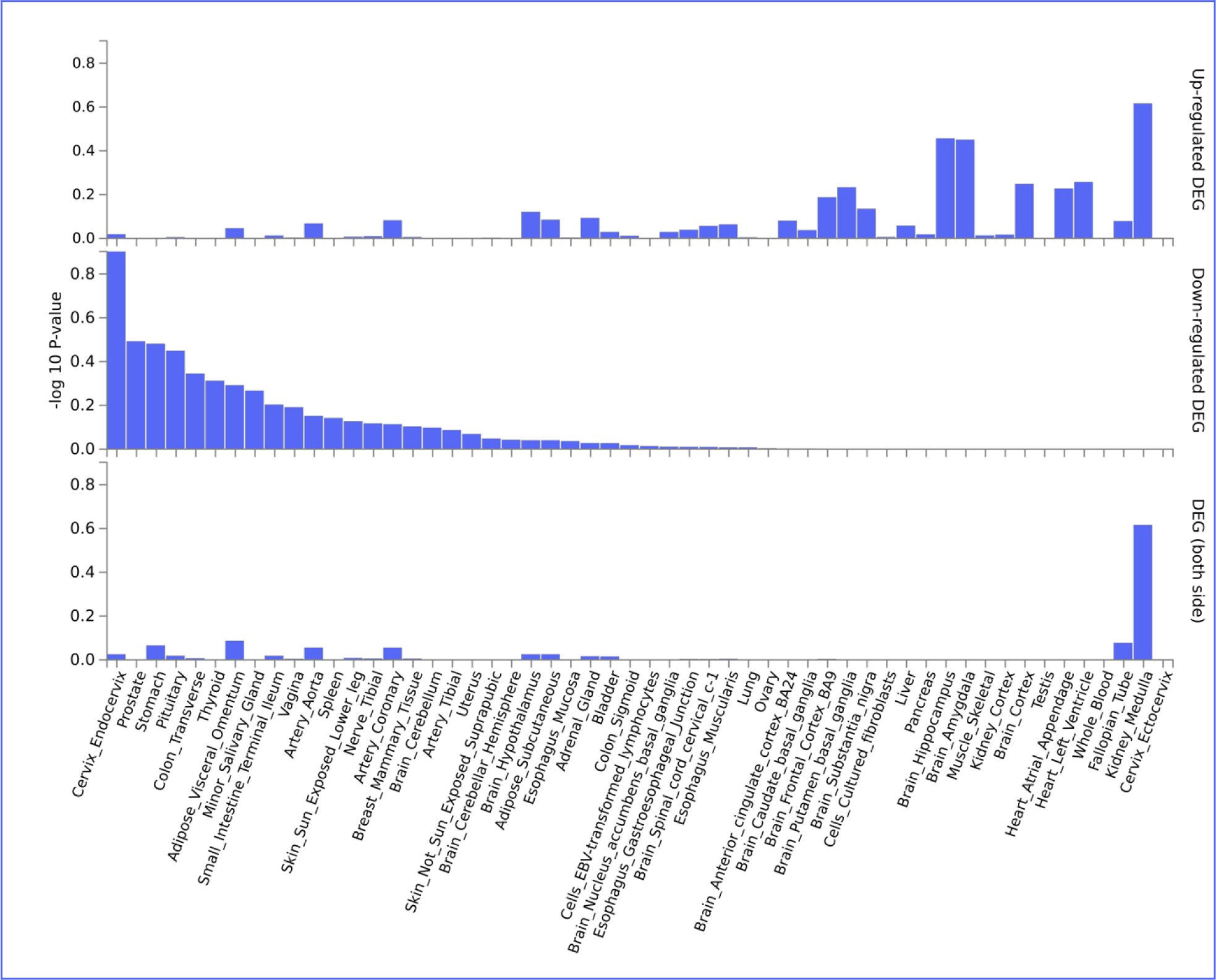
Enriched differentially expressed gene (DEG) sets based on RNA-seq data from 54 tissue types from GTEx v8 ((GTEx Consortium, 2020). Many of the genes mapped to suggestive associations are differentially expressed in endocervical tissue, which is composed of the columnar epithelial cells that line the cervical canal and internal os. The cervical tissue samples includedGTEx V8 were collected from 10 non-pregnant donors, and do not reflect gene expression changes that occur during pregnancy.

## Notes

### Competing Interest Statement

The authors have declared no competing interest.

### Author Declarations

Participants were enrolled under the protocols for Biological Markers of Disease in the Prediction of Preterm Delivery and for Preeclampsia and Intra-Uterine Growth Restriction: A Longitudinal Study (WSU IRB#110605MP2F and NICHD/NIH# OH97-CH-N067) between 2005 to 2017 at the Center for Advanced Obstetrical Care and Research (CAOCR) at Hutzel Womens Hospital. The Institutional Review Boards of Wayne State University and the Eunice Kennedy Shriver National Institute of Child Health and Human Development (NICHD)/National Institutes of Health (NIH)/U.S. Department of Health and Human Services (DHHS) (Detroit MI USA) approved the study. All participants provided written informed consent for the collection of cervical length data and blood samples for future genetic research studies.

